# Estimating (stage-)sojourn time for multiple cancer-sites: literature review and structured elicitation of expert beliefs

**DOI:** 10.64898/2026.06.26.26355688

**Authors:** Dina Jankovic, Stephen Palmer, Matthew E.J. Callister, Georgios Lyratzopoulos, Sofia Dias, Nicky J. Welton, Katherine Payne, Marta O. Soares

## Abstract

Preclinical cancer sojourn time—defined here as the duration a cancer is undetected but detectable—is important for understanding disease progression and evaluating screening policies. This study aims to robustly characterise empirical evidence and existing knowledge over mean sojourn times across 21 stageable tumour sites, including stage-specific preclinical cancer sojourn times and the sojourn time of circulating tumour DNA (ctDNA)-positive cancers.

We updated an existing literature review through to February 2025 to extract population-level empirical sojourn time estimates derived from mathematical models of primary screening data. To synthesise this heterogeneous literature, quantify uncertainty, and obtain estimates for cancer-sites lacking empirical evidence, we conducted a formal Structured Expert Elicitation involving 15 clinical experts. The elicitation was grounded on the literature review results, supplemented by an evidence dossier that included survival data and outcomes from relevant ctDNA cancer studies.

The literature review revealed heterogeneity in existing literature, which focused on a small subset of screened cancers (e.g., breast, cervical, colorectal). The elicitation successfully generated comprehensive probability distributions of overall mean sojourn times for all 21 cancer-sites (representing the site of tumour origin), as well as stage-specific sojourn times and overall sojourn times for ctDNA-positive cancers across 14 cancer-sites.

This study used robust methodology to quantitatively describe existing evidence and experts’ beliefs on the sojourn time of multiple cancer-sites, also describing uncertainty. Such estimates are important for future evaluations of the clinical impact, potential for overdiagnosis and subsequent cost-effectiveness of emerging screening technologies, including multi-cancer detection tests.

## 1. Introduction

The duration of preclinical cancer, or sojourn time, can provide an understanding of cancer progression, inform the potential impact of screening including immediate benefits (e.g. detection) and harms (e.g. overdiagnosis), and support the extrapolations required to evaluate alternative screening options including their long-term consequences.[1]

Sojourn time is an inherently unobservable quantity and cannot be directly estimated. It can, however, be inferred from primary screening data using natural history of disease progression models[2,3]. While multiple definitions of sojourn time exist in the literature, we define it here as the duration of undetected but detectable cancer, representing the precise quantities that can be inferred from primary screening data. To capture disease progression more granularly, overall sojourn time can be broken down by cancer stage to define stage-specific sojourn times. These reflect the duration of time a cancer spends in a given preclinical stage before progressing to a more advanced preclinical stage or being detected. Contextual definitions are provided in Box 1.

**Box 1:**
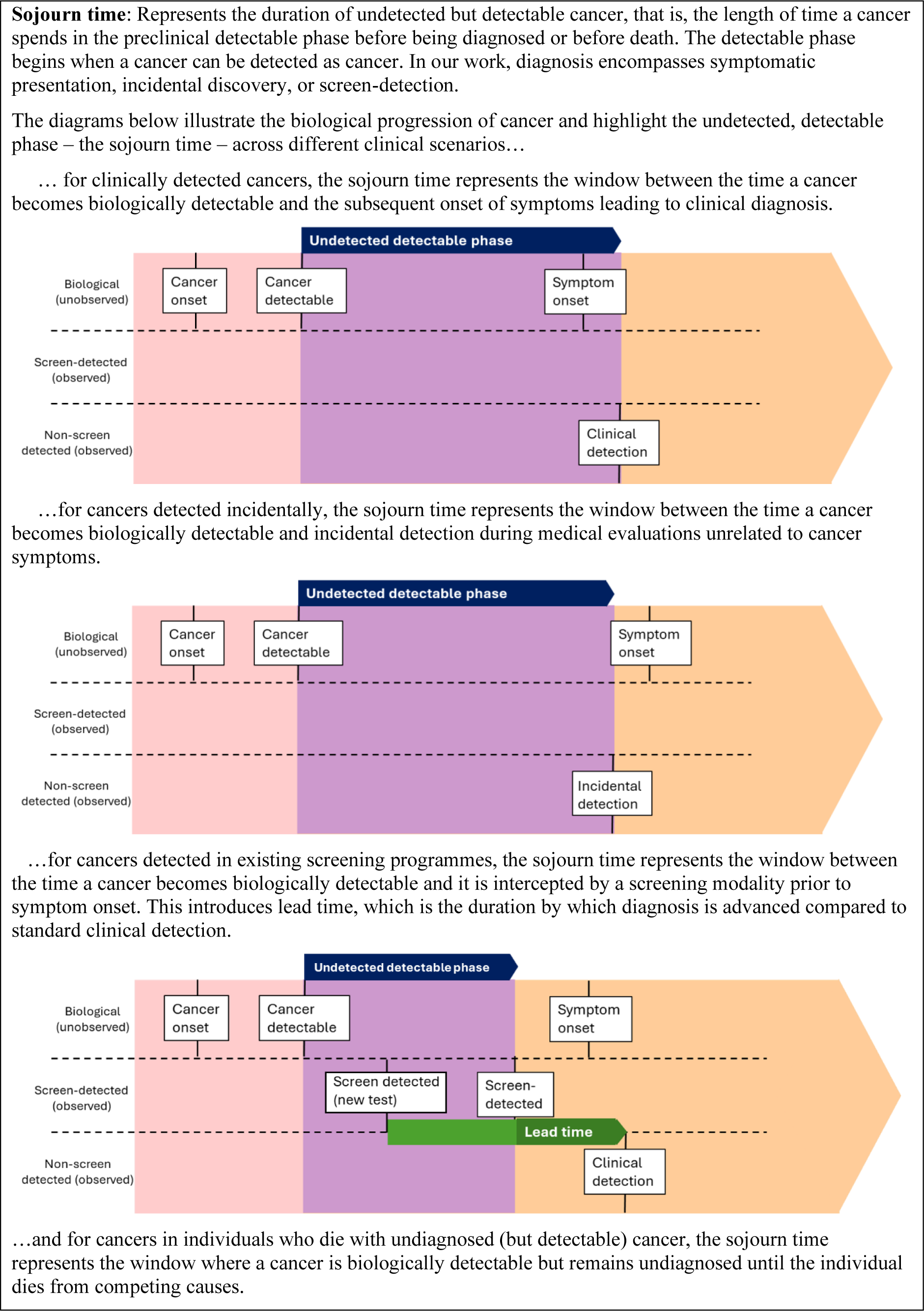
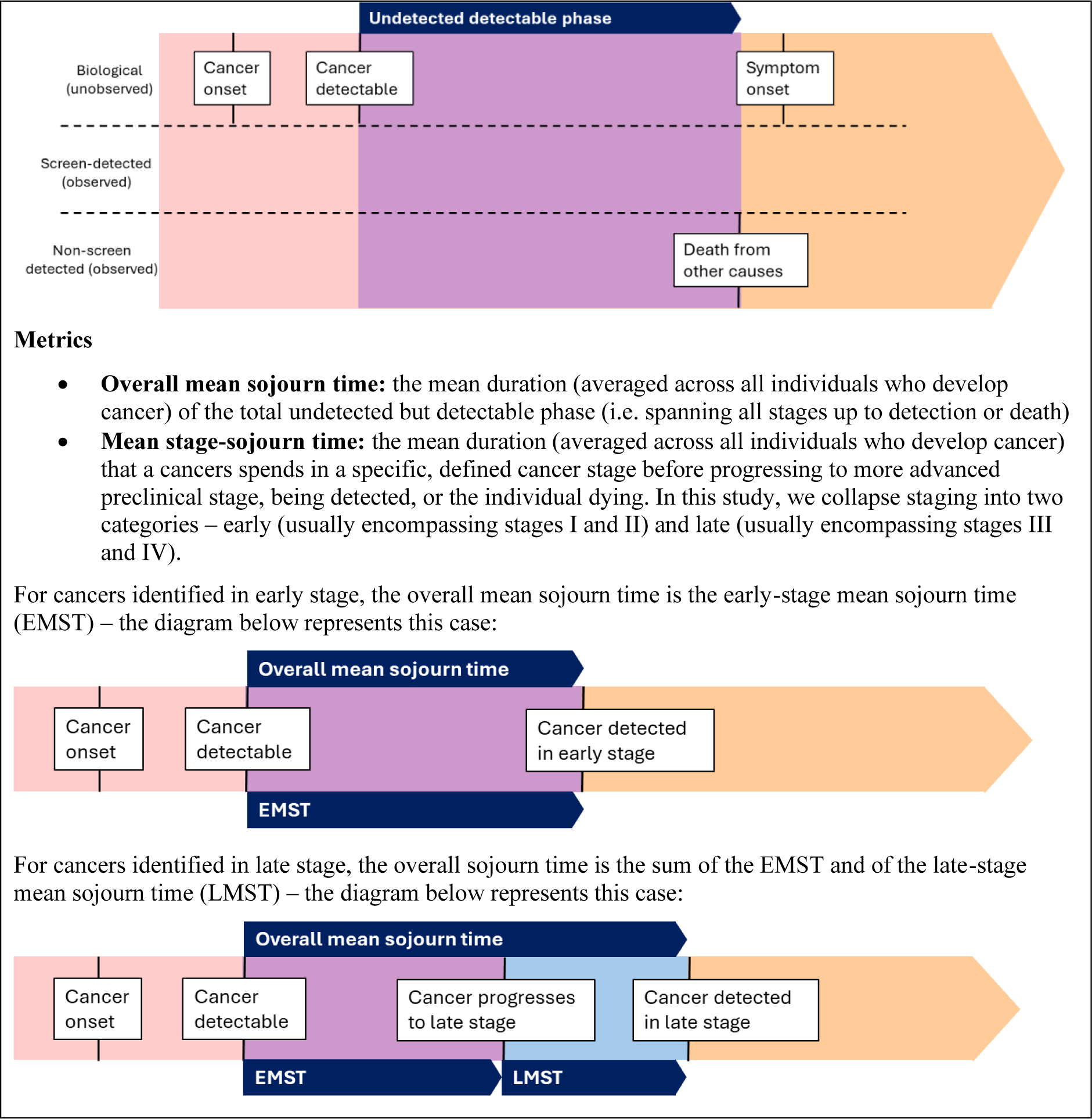
Definitions of (stage-)sojourn time used in this paper.

An important use of evidence regarding mean (stage-)sojourn times is to support evaluations of screening and their optimal specifications. While relevant across traditional (single-)cancer screening evaluations, these estimates are especially important for evaluating multi-cancer tests (MCT) which detect biological signals for a wide range of cancers from a single blood draw.

Because it is unfeasible to design clinical trials for MCTs to be statistically powered to inform outcomes for individual cancer-sites, cancer-specific (stage-)sojourn times are crucial in informing how any potential stage-shifts observed in screening trial data are likely to distribute across specific tumour of origin sites (cancer-site). To do so, sojourn times need to be integrated on a natural history model (defined in the absence of clinical trial data) of cancer-stage progression. To date, such models have relied heavily on a mixture of selective and sparse literature and unstandardised expert input to inform these parameters.[4–9] Estimates informed by experts[4–6,9] were elicited in a study[10] where sojourn times were not the primary focus, the definition of sojourn times was not clear, experts were provided limited background information on which to base their estimates, and the estimates did not capture experts’ uncertainty. Estimates derived from the literature[7,8] have relied on selected (single) estimates with limited justification provided for their selection or their generalisability to other cancer-sites included in the models.

In this paper, we aim to systematically describe current evidence and knowledge regarding mean sojourn times across 21 cancer-sites, for all cancers (overall and stage-specific) and for ctDNA cancers (only overall), while explicitly quantifying the uncertainty surrounding these estimates. To achieve this, we first updated a literature review of the existing literature from primary screening studies to identify empirical estimates of sojourn times. We then supplemented this with a formal Structured Expert Elicitation (SEE) [11], grounded on the empirical findings of the review. The elicitation helps interpret and summarise the existing empirical estimates, which are expected to be heterogeneous, as well as to document and justify assumptions concerning the generalisability of these estimates to select cancer-sites (or groupings) without existing empirical evidence. Because we intend for our estimates to inform future MCT evaluations, we specifically elicited mean sojourn times for cancers identified via circulating tumour DNA (ctDNA), the methodology used by the Galleri® (GRAIL) blood test. Cancers with a positive ctDNA signal are hypothesised to have shorter sojourn times[12–15]. We use the elicitation to document and justify experts’ beliefs on how this sojourn time compares to that of all cancers.

The remainder of the paper is structured as follows: Section 2 describes the literature review (2.1) and elicitation (2.2) methodologies; Section 3 presents the empirical estimates from the review (3.1) and from the elicitation (3.2); and Section 4 summarises our findings and discusses their implications for policy and future evaluations.

## 2. Methods

### 2.1. Literature review

We updated a published literature review [2] that aimed to identify empirical estimates of mean sojourn time inferred from mathematical models applied to the analyses of primary screening data. Further details on the methodology are in Supplementary files S1. We further excluded papers that: did not report population-level estimates of (stage-)mean sojourn time, that reported estimates calculated from prevalence-incidence ratios only (further detail below), and that relied solely on empirical data that relates to screening tests with poor sensitivity for that cancer-site, including studies examining the use of X-rays for lung cancer screening.

We extracted information on the study, screening dataset, mathematical model and sojourn time estimate. Mathematical models included: Maximum Likelihood Estimation (MLE), where a likelihood function built around a specific disease natural history models is maximised; Expectation-Maximization Algorithm (EMA), an iterative optimisation variation of MLE, used to handle latent data, in this case, the time a cancer enters its preclinical phase; Regression of Observed on Expected (ROE), an alternative regression-based technique that minimises the discrepancy between empirical screening observations and the expected outcomes of a natural history of disease model defined on sojourn time; and Bayesian models, which also define a natural history of disease model but use a distinct inferential approach that structurally integrates the primary data with prior probability distributions, allowing uncertainty to be integrated more seamlessly than other methods. We excluded prevalence-incidence ratio calculations, which arithmetically derives sojourn times strictly based on the ratio of preclinical cancer prevalence to clinical incidence. This method relies on stronger underlying assumptions, including that the screening test are 100% sensitive, and is known to be less accurate.[2,16]

We summarised all estimates of interest (point estimate and associated uncertainty, where available) in each study, grouped by dataset and modelling approaches used to derive them (MLE, EMA, ROE or Bayesian).

### 2.2. Structured Expert Elicitation (SEE)

In the elicitation, we aimed to inform, by cancer-site:

- Overall mean sojourn time, and level of variation across individuals;
- Mean early and late stage-sojourn times, elicited separately for cancers diagnosed in early-stage and in late stage; and
- Overall mean sojourn time for ctDNA positive cancers, operationally defined as those that would have yielded a positive ctDNA signal via blood-based MCTs, specifically to the Galleri test.

The elicitation was designed using formal established methods[11,17,18], employed to support experts to consider all available evidence carefully, to minimise common biases, and to capture experts’ beliefs accurately. It was protocolled (Supplementary file S2). Importantly, individuals were asked to express their beliefs over a plausible range of values rather than a single value, being able to express uncertainty in their knowledge. They were therefore able to express full uncertainty if they have no information on the quantities of interest.

Before answering questions, experts were trained, provided with definitions of sojourn times (as per Box 1), and with a summary of evidence sources and evidence gaps and, for each cancer-site, which included: empirical estimates of overall mean sojourn times informed by the review (Section 2.1); five-year survival and staging at diagnosis in England [19,20]; information on existing cancer screening programmes (breast, colorectal, lung and cervical); a summary of evidence on ctDNA cancers, including Galleri’s test sensitivity from the CCGA3 study[21], overall cancer survival for cancers detected vs. not detected by Galleri[14,15], and ctDNA detection rate 1, 2 and 3 years prior to conventional cancer diagnosis [12,13]. The training slides, and the evidence dossier and example elicitation questions are in the Supplementary files S3 and S4, respectively.

We recruited practicing clinicians in the UK National Health Service who considered themselves to have good knowledge of screening research. Experts were purposely selected, e.g. through publications (relating to sojourn times or ctDNA cancers), participation on relevant boards (e.g. the UK National Screening Committee) and on recommendation by peers and professional bodies, namely UK Collaborative for Cancer Clinical Research (UKCCCR). The sample included “specialists” (typically surgeons, oncologists or clinicians in the oncology diagnostic pathway who specialise in one or a subset of cancer-sites) and “generalists” (e.g. general oncologists and GPs) who specialise in early cancer detection rather than any specific cancer-site. While specialists could provide more precise estimates of sojourn times in cancer-sites within their specialty, generalists may have a better understanding of relative overall mean sojourn time between different cancer-sites. In the elicitation, specialists only provided answers for cancer-sites within their speciality, while generalists were asked about all 21cancer-sites. We ensured at least four experts elicited for each of the 11 key cancer-sites.

The elicited quantities are summarised in detail Table 1, as well as the rationale for the chosen quantities and how they relate to the above parameters we ultimately aimed to inform. Further detail on the derivations is presented in Supplementary files S5.

**Table 1.** Elicited quantities and rationale.

| Question number | Quantity | Rationale |
| --- | --- | --- |
| Q1a | Overall mean sojourn time for clinically detected cancers (elicited with uncertainty) | Includes both ctDNA positive and negative cancers combined. +<br>It is the only sojourn time parameter with empirical estimates. Elicitation will capture experts' beliefs about the validity and generalisability of the empirical estimates. |
| Q1b | Overall mean sojourn time including screen-detection for cancer-sites with a screening programme (breast, colorectal, cervical and lung) (elicited with uncertainty) | Informs overall mean sojourn time of all cancers detected in the UK, in absence of the Galleri test. Unlike Q1a, the sojourn time estimate considers the lead time of screen-detected cancers. |
| Q1c | 25th percentile of overall sojourn time for all cancers detected in the UK (point estimate only elicited)<br>This is elicited conditioned on assuming the mode of Q1a/Q1b** is the true overall mean sojourn time | Describes heterogeneity in overall sojourn time. This is used internally to:<br>• validate the use of the Exponential distribution to describe overall sojourn time, or to fit an alternative distribution.<br>• predict mean sojourn time for the x% of the cancers with the shortest sojourn time, using the fitted distribution.* |
| Q2a | Overall mean sojourn time of cancers diagnosed in early stage (point estimate only elicited)<br>This is elicited conditioned on assuming the mode of Q1a/Q1b** is the true overall mean sojourn time | This is equivalent to the early mean sojourn time of cancers diagnosed in early stage. This is used internally to:<br>• infer the overall mean sojourn time of cancers diagnosed in late stage using the % diagnosed in late stage and the overall mean sojourn time. |
| Q2b | Early-stage mean sojourn time for cancers diagnosed in late stage (point estimate only elicited)<br>This is elicited conditional on overall mean sojourn time for cancers diagnosed in late stage inferred in Q2a | Informs late-stage mean sojourn time (overall mean sojourn time minus early-stage mean sojourn time) for cancers diagnosed in late stage. |
| Q3 | Overall mean sojourn time for ctDNA cancers + (elicited with uncertainty)<br>This is elicited conditioned on assuming the mode of Q1a/Q1b** is the true overall mean sojourn time | Qualitative questions to establish whether ctDNA positive cancers are those with shortest sojourn time and, if so, whether the prediction in Q1c is valid. If none of the previous, the expert is asked to elicit directly with uncertainty. |

+ ctDNA positive cancers were defined as those that would have yielded a positive ctDNA signal via blood-based MCTs, specifically to the Galleri test.
\* x represents the proportion of cancers that are ctDNA positive, estimated from the Galleri test sensitivity in the CCGA3 study (Klein et al., 2021)
\*\* For breast, colorectal and cervical cancers we used the mode of overall mean sojourn time for clinically and screen-detected cancers (Q1b) to represent current practice in absence of the Galleri test. For lung cancer we used the mode OMST without screening (Q1a) as the screening programme is still being rolled out and its impact on cancer detection has not yet been observed.

The elicitation was conducted in live, remote and individual sessions. facilitated by an experienced researcher (DJ). Elicitation questions were answered using a bespoke online tool designed in RShiny available at https://shiny.york.ac.uk/cancer_sojourn_time/. Informed consent was sought from each participant. The consent form and protocol were approved by the University of York’s Health Sciences’ Research Governance Committee (HSRGC/2025/699/E) on the 7 July 2025, prior to the conduct of the exercise.

Responses from individual experts were pooled mathematically and group consensus was not sought[11]. The individually elicited quantities were linearly pooled, a method that preserves the individual judgements in the collective (pooled) judgement[22]. In pooling, equal weights across experts were used. To represent uncertainty, we fitted a distribution to the pooled elicited summaries using the *fitdist* function of the SHELF package in R[23]. Lognormal and Gamma distributions (both having strictly positive support) were considered, and the best fitting (lowest sum of squared errors between elicited and fitted probabilities) was selected. The analysis code with a sample of synthetic responses is available at https://github.com/jankovicd/cancer_sojourn_times_analysis.

To establish face validity of the findings of our study, the individual experts were sent displays of the results of the elicitation (a draft of the results section of this paper) and were asked whether the results broadly reflected their expectations for the cancer-sites they elicited.

## 3. Results

### 3.1. Literature review

From updating the review by Geurts et al. (2022)[2], we identified and screened 332 unique records, identifying 13 new publications with estimates of mean sojourn time (see Supplementary files S1 for details). Combined with the original review, of which we used 15 studies, we included a total of 26 studies across six cancer-sites. Table 2 shows there is significant variation in the level of evidence across cancer-sites, with the most evidence existing for breast cancer.

**Table 2.**
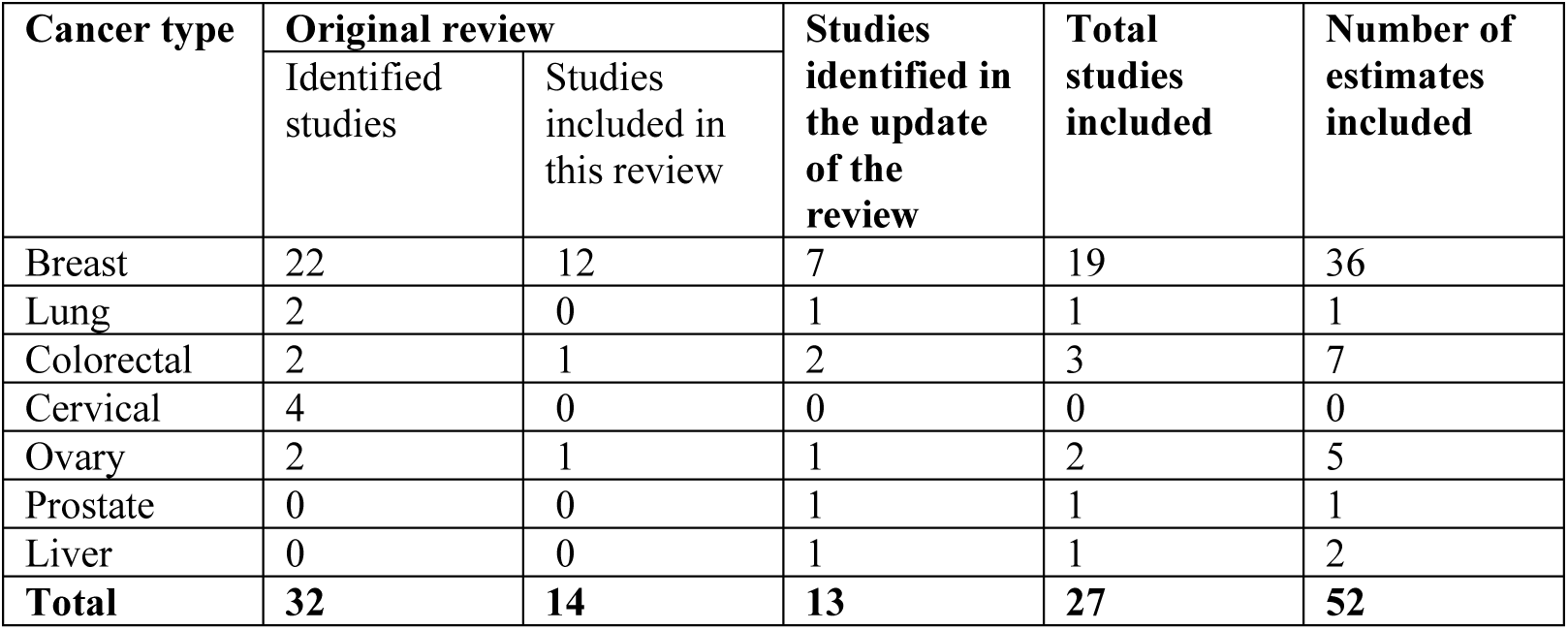
Number of studies identified in the updated literature review, by cancer type.

Figure 1 presents a forest plot summarising overall mean sojourn time estimates from each study (no study reported stage-specific estimates), grouped by datasets and by modelling approaches used to derive them. The evidence in Figure 1 shows that, within breast cancer, there is significant variation in overall mean sojourn time estimates, arising from the dataset (for example, estimates from the HIP trial for example from [24] seem generally shorter than those obtained from other breast cancer datasets), from the year of the data (for example, estimates across the Nijmegen cohorts generally increase over time[25]), from the method of analysis (for an example, see estimates from any one of the Nijmegen cohorts[25]), and from assumptions underlying the analysis (for example, see the estimates in [26] which apply the same method, MLE, under different assumption).

**Figure 1:**
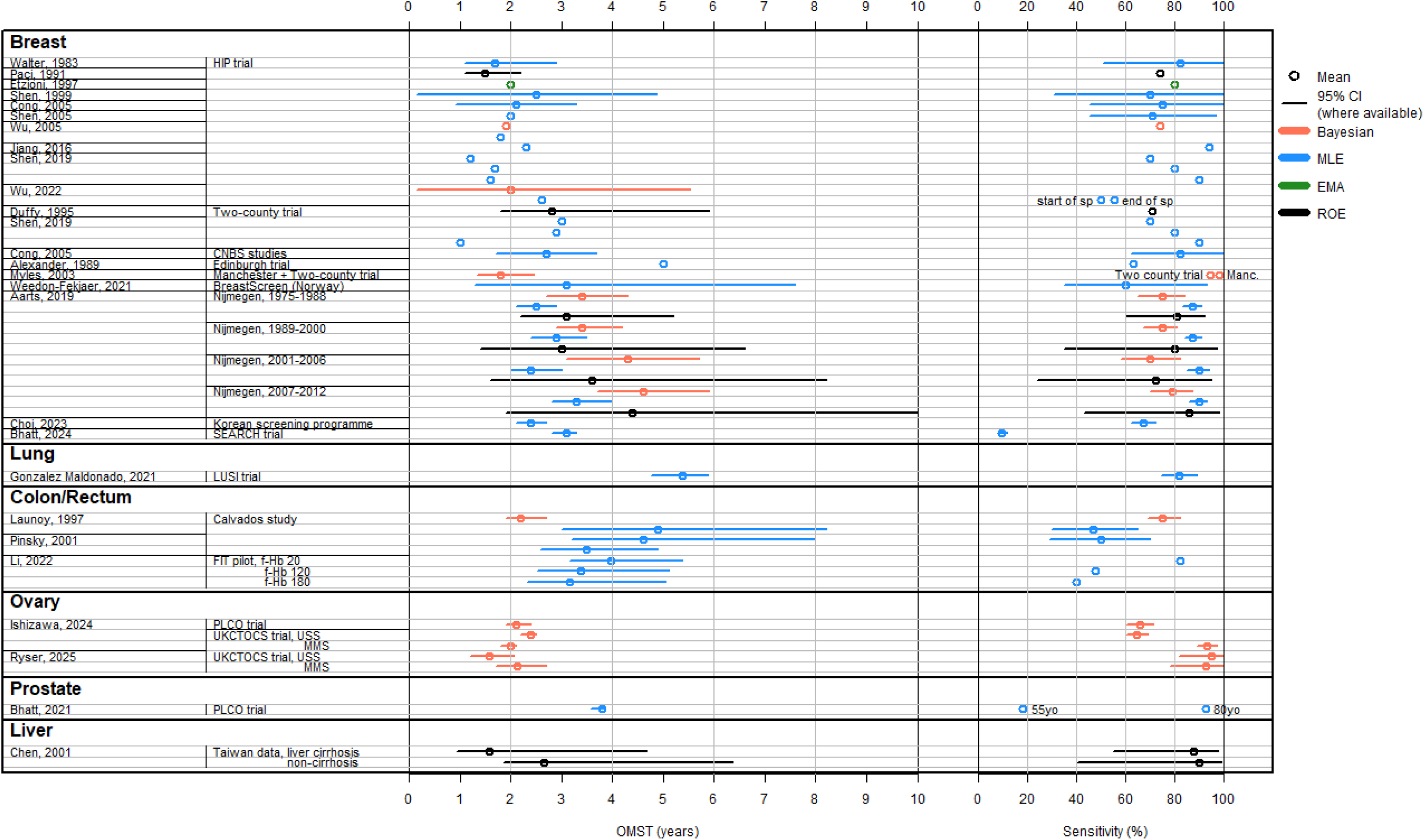
Forest plot of estimates of OMST and test sensitivity in the literature by cancer-site, study, dataset and mathematical estimation method. MLE: Maximum likelihood estimation, EMA: Expectation Maximisation Algorithm, ROE: Regression of Observed on Expected. Estimates are presented with 95% confidence/credible intervals, except where these were not available in the original publication. Multiple estimates in the same study, using the same dataset and the same mathematical estimation method represent analyses using different modelling assumptions, which were out of scope for our work and were therefore not described.

Figure 1 shows that the estimate available for lung cancer is the longest across cancer-sites (point estimate above 5 years), which may be because these cancers lie mostly undetected until late-stage disease. Ovary presents short overall mean sojourn times, and homogenous estimates across studies and datasets, which may be a result of the fact that this cancer-site is itself relatively homogeneous.

### 3.2. Elicitation results

We elicited from 15 experts: 12 specialists (five surgeons, two oncologists and five consultants in medicine - gastroenterology, respiratory medicine and gynaecology) and 3 generalists (professors in either epidemiology or primary care diagnostics). A breakdown of experts by cancer-site is reported in Supplementary files S6, Table S6.1. On average experts had 19 years of experience in their role (range 8-34), with 14/15 experts holding research positions. Their primary work location was geographically dispersed across eight cities in the England and Northern Ireland (Belfast, Birmingham, Cambridge, Exeter, Leeds, Liverpool, London, Nottingham).

In total, experts quantitatively elicited 55 overall mean sojourn times (including for all cancers and for ctDNA cancers separately), 55 estimates of variability, and 42 mean stage-sojourn times. In addition, 29 overall mean sojourn times were elicited qualitatively, by asking experts to select one or more of the previously elicited cancer-site they are most similar to. The number of experts who elicited for each cancer-site ranged from two (urothelial tract) to six (lung, pancreas), with at least one specialist and three generalists eliciting for the 11 cancer-sites prioritised in the elicitation.

On visual inspection, we found no consistent trends in the direction of bias or level of uncertainty that individual experts expressed compared to the pooled estimates. In this section we present pooled estimates for specialist, generalist and all experts combined, while in Supplementary files S6 we present anonymised individual estimates.

#### Overall mean sojourn time for all cancers and variability

The pooled expert estimates for overall mean sojourn time are shown in Figure 2, presenting boxplots with the median and interquartile range, overlaid with a forest plot representing 95% credible intervals. For individual estimates see Figure S6.1 in Supplementary file S6. Figure 2 shows there is some variation between cancer-sites. Prostate cancer has the longest overall mean sojourn time (pooled median of 6 years for all experts combined), and ovarian cancer the shortest (2 years for all experts combined). Median estimates for ovarian cancer are comparable to liver and pancreatic cancers. This is likely influenced by the precise empiric estimates from studies (Figure 1) that experts highlighted as good quality studies during the elicitation.

**Figure 2.**
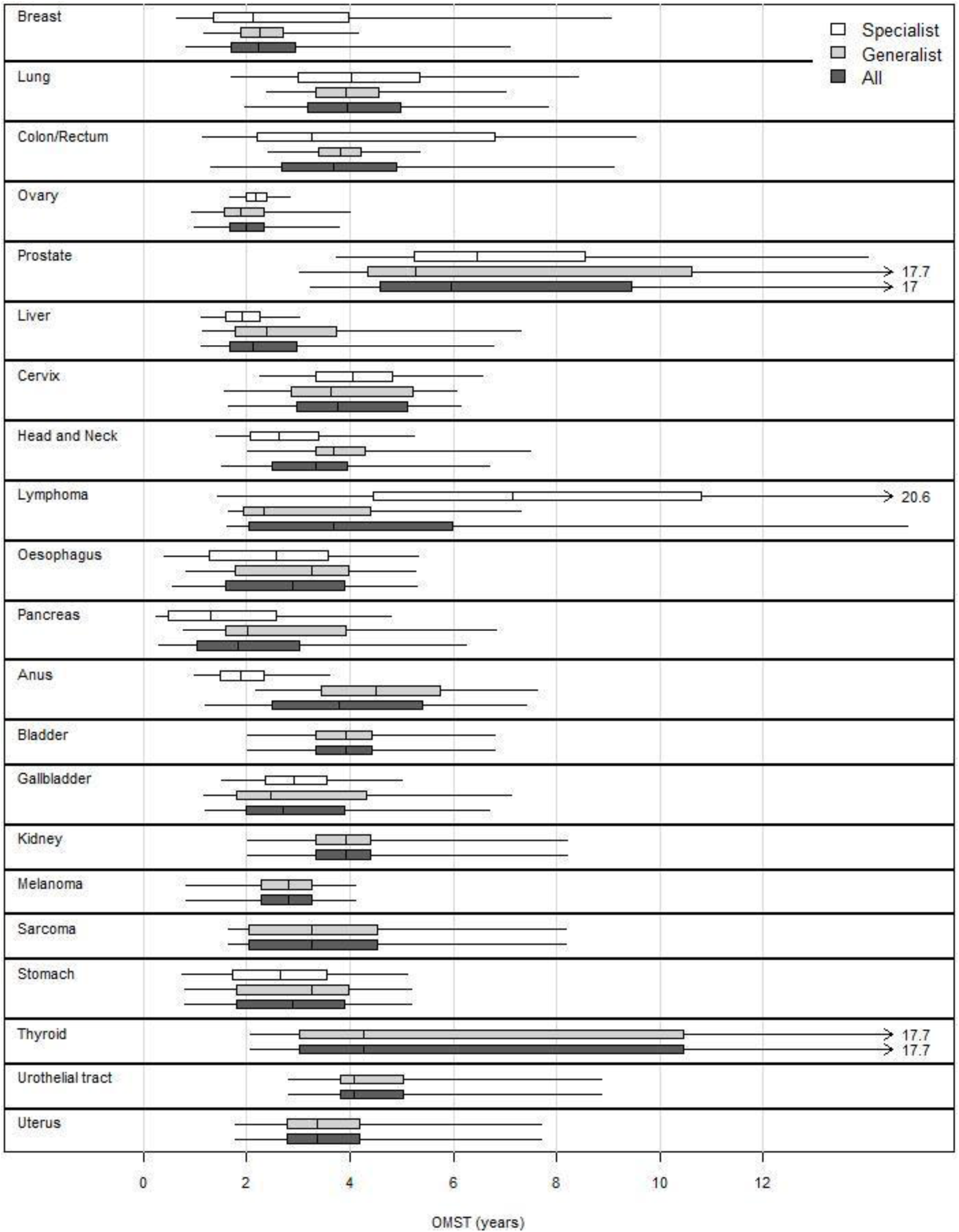
Pooled estimates on overall mean sojourn time, by cancer-site (includes screen-detected cancers for sites with fully implemented screening programmes: breast, cervical and colorectal). Boxplots show the median, interquartile range and 95% credible intervals.

We see no consistent differences in results between specialists’ and generalists’ estimates. We hereafter only report estimates pooled across all experts.

For cancer-sites with screening programmes, experts consistently indicated that the inclusion of screen-detected cancers shortened the overall mean sojourn time (Supplementary file S6, Figure S6.2). This is expected, as screening intercepts cancers early, although one expert expressed high uncertainty around their estimates stating that screen-detected cancers, particularly breast cancers, were likely to include indolent cancers with very long overall mean sojourn time and therefore could lead to higher estimates even if those cancers are technically being detected earlier.

The degree of agreement between the available empirical estimates in Figure 1 (available to expert at the time of elicitation) and the elicited estimates in Figure 2 varied between cancer-sites. Experts’ estimates for breast, colorectal and liver cancers are reflective of the available evidence, presenting wide confidence intervals and medians close to empirical point estimates. Elicited estimates for ovarian cancer are also reflective of empirical estimates, with central estimates close to the empirical estimates but with relatively narrow confidence intervals likely influenced by the precise empirical estimates from studies that experts highlighted as good quality studies during the elicitation.

The elicited estimate for lung cancer was lower and more uncertain than the one available empiric estimate. This was, however, elicited consistently across experts, and justified by the small number of empirical estimates with uncertain generalisability, and by the estimates for other cancer-sites.

Elicited estimates for prostate cancer were substantially higher and more uncertain than the one empirical estimate. This was consistent across all experts, who believed that a significant proportion of prostate cancers are very slow progressing and remain undetected for long periods of time.

We also elicited variability (results in Supplementary File Table S6.2) by asking experts for the 25^th^ percentile of the sojourn time (i.e. the maximum sojourn time for the 25% of individuals with the shortest sojourn time). Of the 54 elicited 25th percentiles, only one was consistent with the exponential distribution; for the remaining, the logNormal distribution was therefore used.

#### Mean stage-sojourn times

Derived mean stage-sojourn times are shown in Figure 3. The figure presents, for each cancer-site, overall mean sojourn time for all cancers, for those detected in early stage and for those detected in late stage. The results suggest that the relationship between overall mean sojourn time for cancers detected in early stages and in late stages differed across cancer types. In three cancer-sites (breast, lymphoma and pancreas) overall mean sojourn time was judged to be similar between cancers diagnosed in early and late stages; in four cancer-sites (colorectal, liver, head and neck and bladder) overall mean sojourn time was judged to be shorter for cancers diagnosed in early stages than in late stages, and in the remaining seven cancer-sites overall mean sojourn time was judged to be longer for cancers diagnosed in early stages than in late stages.

**Figure 3.**
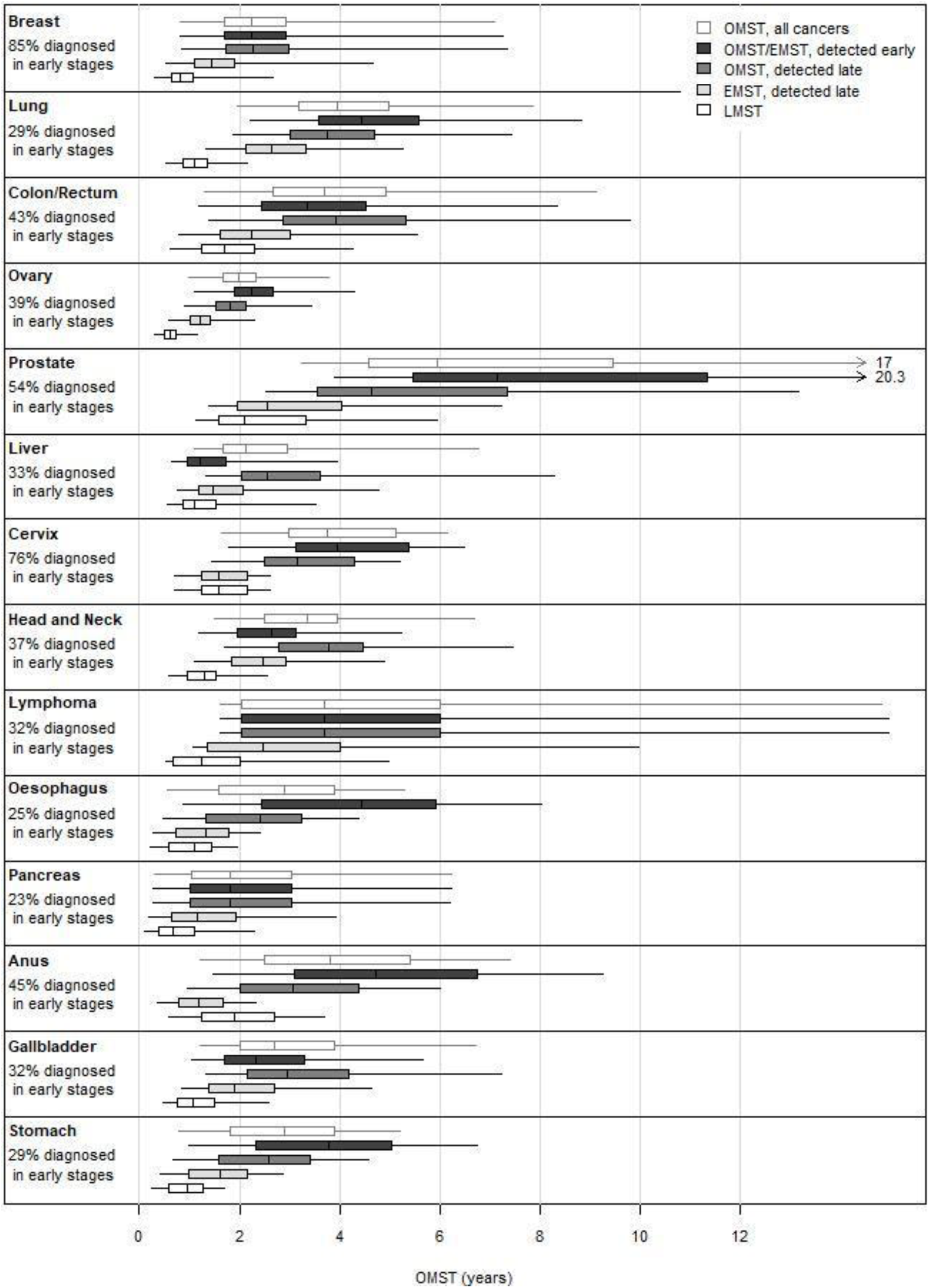
Derived mean stage-sojourn time for all cancers, by cancer-site. Boxplots show the median, interquartile range and 95% credible intervals. OMST: overall mean sojourn time. EMST: early-stage mean sojourn time. LMST: late-stage mean sojourn time.

Figure 3 also presents the breakdown of overall mean sojourn time by stage (early and late) for cancers diagnosed in late stages. Cancers diagnosed in late stages were judged to progress faster (shorter early stage mean sojourn time) for all cancer-sites except the liver, where all three experts suggested that early stage sojourn time is either equal or longer in cancers diagnosed in late stages (see Supplementary files S6, Figure S6.3). The difference in the duration of early stage between cancer-sites diagnosed early vs late stages varied across cancer-sites, from 1% difference (ovarian and head and neck cancers) to more than 4 times longer early stage sojourn time if diagnosed in early stages (oesophagus, anus).

For cancer-sites diagnosed in late stage, late stage sojourn time was judged to be shorter than early stage sojourn time for all cancer-sites except the anus, elicited by only one expert. The difference between early and late stage sojourn times varied, from identical (cervical cancer) to 2.4 times longer early stage than late stage (lung cancer).

#### Overall mean sojourn time for ctDNA cancers

Of the 54 responses related to ctDNA cancers (Supplementary files, Table S6.3 and Figure S6.4), 41 (76%) indicated that ctDNA cancers have the shortest overall mean sojourn time of all cancers within that cancer-site. Thirty-four of these (83%) agreed with the overall mean sojourn time point estimate predicted by the statistical distribution chosen (1 Exponential and 33 logNormal); the remaining 20 elicited with uncertainty.

Figure 4 shows the predicted overall mean sojourn time of ctDNA cancers (shown in Supplementary files, Figure S6.5). For all cancer-sites, experts believed that overall mean sojourn time is shorter in ctDNA cancers than in all cancers, but they vary in the extent of the adjustment. Cancer-sites with lower % of ctDNA cancers (given by Galleri’s test sensitivity) like breast or prostate have a greater adjustment suggesting they are the cancers with the shortest sojourn times. In fact, experts judge that ctDNA positive breast and prostate cancers have a lower median overall mean sojourn time of all cancer-sites.

**Figure 4.**
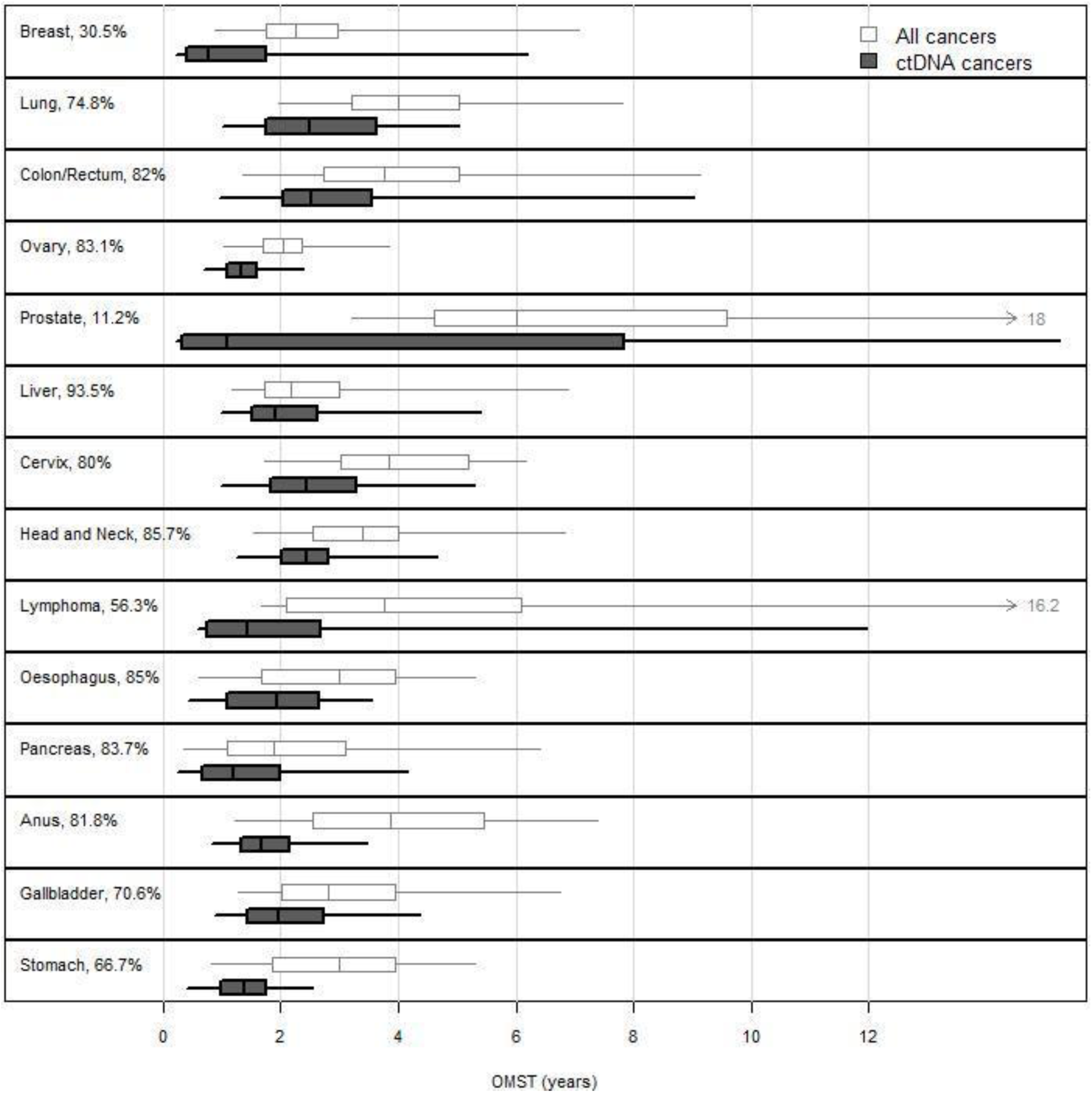
Overall mean sojourn time derived for ctDNA compared to all cancers, by cancer-site.

#### Face validity results

Thirteen of the 15 experts responded to the survey and, of these, 12 responded that the results broadly reflected their expectations for the cancer-sites they elicited. The remaining expert justified that they did not expect to see an overall mean sojourn time for ovarian cancer comparable to breast cancer, and was expecting longer results for prostate cancer, colon/rectum and lung cancer.

## 4. Discussion

Our study presents a comprehensive overview of existing evidence and a structured approach to elicit knowledge regarding mean cancer sojourn times—the period during which a cancer is detectable but remains undiagnosed—across 21 stageable cancer-sites. Using a systematic review, we identified - empirical evidence on overall mean sojourn time from 27 studies covering six cancer-sites. This empirical evidence was then considered by 15 UK clinical experts as part of a structured expert elicitation exercise that generated final estimates for all 21 cancer-sites.

In brief, our findings indicate that overall mean sojourn time varies significantly by cancer-site, with prostate cancer having a long mean sojourn time of approximately 6 years, whereas ovarian, liver, and pancreatic cancers have much shorter durations of around 2 years. A majority of elicitations (31/41, 76%) indicated that early-stage sojourn times are longer in cancers diagnosed at an early stage compared to those diagnosed late, suggesting that cancers diagnosed at late stages may progress inherently faster. In relation ctDNA cancers, our research indicates that these cancers are likely those with the shortest sojourn times within their respective cancer-sites.

Two recent publications [27,28] analysed ctDNA sojourn times using a CPS-3 biobank sample of 1,064 healthy participants who developed defined, stageable, single cancers within three years of enrolment. While preliminary data from earlier conference presentations [12,13] was included in our evidence dossier and informed expert responses, these now-published estimates of median sojourn times range from under 0.5 years (pancreas/esophagus) to just over 2 years (lymphoma/ovary). Comparatively, our study projected fewer cancer types with a median sojourn time below 1 year, with most falling between 1 and 2.5 years.

The elicitation was designed to be as robust as practically possible; it used a structured methodology following existing standards and was designed to minimise common biases and capture expert beliefs as accurately as possible. We purposively selectively recruited individuals who are substantive experts in oncology and early detection research. To ensure high internal validity of the elicitations, specifically that the descriptions elicited reflected as closely as possible the beliefs of experts, the elicitations were conducted by an experienced facilitator (DJ) in one-to-one interviews. All experts were able to complete the exercise, reaching a good understanding of what was being asked and how to record their answers. Written feedback from experts did not highlight any systematic problems with their understanding of the quantities elicited and confirmed the face validity of the results obtained.

Our exercise was based on specific cancer “sites” (i.e. tumour origin in specific organs). Some cancer-sites present histological heterogeneity (for example, breast, ovary, lymphoma, oesophageal, and lung), while others are dominated by a single cancer type (e.g., adenocarcinomas constitute the great majority of colon/rectal cancers). Cancers with morphological heterogeneity may still show a similar prognosis and therefore a similar sojourn time, in which case it remains appropriate to elicit sojourn time at the ‘cancer-site’ level. This appears to be the case for lung, oesophageal and liver cancers.

Where different cancer types within the same organ present different prognosis, the sojourn time may also differ significantly. Examples of this may be breast and ovarian cancers and lymphoma. While pancreatic cancer is chiefly represented by one type (adenocarcinoma), the small proportion that are not adenocarcinoma present significantly better prognosis. While here we elicited the mean sojourn time across morphological types, it would be important for further research to examine sojourn time by morphological type for those expected to have distinct prognosis.

In conclusion, the estimates of sojourn times generated in our study contribute to the understanding (and description) of pre-clinical cancer progression and can be used in support of policy and practice decisions about diagnosis, detection and screening in oncology. A specific example consists in using these estimates, as priors or directly, to inform the natural history component of screening models. By providing estimates across a large number of cancer-sites, our work is of particular relevance for the evaluation of MCT for screening.

## Data Availability

All data produced in the present study are available upon reasonable request to the authors

https://github.com/jankovicd/cancer_sojourn_times_analysis

## Acknowledgements

We would like to thank the 15 experts that took part in the elicitation, which include Willie Hamilton, Laurence Lovat, Keith J. Roberts, Nora Pashayan, Shreerang Bhide, Matthew E.J. Callister, Hashim U. Ahmed, Georgios Lyratzopoulos, Emma O’Dowd, Stephen P Pereira, Usha Menon, Robert P. Jones and Jeffery Smith, which agreed to be named here, and another two additional anonymous experts.

## Funding disclaimer

This project is funded by NHS England.

Katherine Payne is supported by the National Institute for Health and Care Research (NIHR) Manchester Biomedical Research Centre (BRC) (NIHR203308). Katherine Payne is a NIHR Senior Investigator.

The views expressed are those of the authors and not necessarily those of the funders.

