## Supplemental Files for "Estimating (stage-)sojourn time for multiple cancer-sites: literature review and structured elicitation of expert beliefs"

#### Supplementary information

#### S1. Updated literature review: methods and results

##### S1.1 Methods

We updated a published literature review, Geurts et al. (2022), that aimed to identify empirical estimates of mean sojourn time inferred from mathematical models applied to the analyses of primary screening data.

The original search strategy was composed of 3 parts—(1) cancer; (2) screening; and (3) PCDP, sojourn time, or lead time (reproduced in Table S1.1 below) —and was performed in PubMed and Embase on February 8, 2018. This was re-run, with the addition of date limits set to the period following this existing search (after 8.02.2018), and up to 02.2025. The original search included primary studies in English or Dutch; the updated included studies in English only.

**Table S1.1.** Embase and PubMed search strategy with used search terms; reproduced from Geurts et al. (2022).

| Database | Part of the search | Search terms (Mesh and free text) |
|---|---|---|
| **Embase** | **Cancer** | "Neoplasms"[Mesh] OR cancer.ti,ab. OR cancers.ti,ab. OR carcinoma.ti,ab. OR neoplasm.ti,ab. OR neoplasms.ti,ab. OR .ti,ab. OR s.ti,ab. OR tumour.ti,ab. OR tumours.ti,ab. OR neoplasia.ti,ab. |
| | **Screening** | "Mass Screening"[Mesh] OR screening.ti,ab. OR screenings.ti,ab. OR screen.ti,ab. OR screens.ti,ab. OR screened.ti,ab. OR screen-detected.ti,ab. OR screen detected.ti,ab. OR "Early Detection of Cancer"[Mesh] OR early Detection.ti,ab. OR early diagnosis.ti,ab. |
| | **Preclinical detectable phase (sojourn time)/lead time*)** | lead time.ti,ab. OR lead-time.ti,ab. OR sojourn time.ti,ab. OR sojourn-time.ti,ab. OR Pre-clinical disease state.ti,ab. OR preclinical disease state.ti,ab. OR Pre-clinical detectable phase.ti,ab. OR preclinical detectable phase.ti,ab. OR detectable preclinical phase.ti,ab. OR preclinical duration.ti,ab. OR preclinical detectable disease state.ti,ab. |
| **PubMed** | **Cancer** | "Neoplasms"[Mesh] OR cancer[tiab] OR cancers[tiab] OR carcinoma[tiab] OR neoplasm[tiab] OR neoplasms[tiab] OR [tiab] OR s[tiab] OR tumour[tiab] OR tumours[tiab] OR neoplasia[tiab] |
| | **Screening** | "Mass Screening"[Mesh] OR screening[tiab] OR screenings[tiab] OR screen[tiab] OR screens[tiab] OR screened[tiab] OR screen-detected[tiab] OR screen detected[tiab] OR "Early Detection of Cancer"[Mesh] OR early Detection[tiab] OR early diagnosis[tiab] |
| | **Preclinical detectable phase (sojourn time)/lead time*)** | lead time[tiab] OR lead-time[tiab] OR sojourn time[tiab] OR sojourn-time[tiab] OR Pre-clinical disease state[tiab] OR preclinical disease state[tiab] OR Pre-clinical detectable phase[tiab] OR preclinical detectable phase[tiab] OR detectable preclinical phase[tiab] OR preclinical duration[tiab] OR preclinical detectable disease state[tiab] |

* Lead time was included in the search strategy as we found out that this term is sometimes mixed-up with the preclinical detectable phase duration.

We supplemented our electronic database search by consulting our advisors, content experts in early detection research. These experts (GL and MC) were asked to identify relevant published or unpublished studies that might have been missed by the initial database search strategy.

As with the original search, we selected studies that described new or adapted mathematical models designed to estimate the duration of the sojourn time (referred to as preclinical detectable phase, PCDP, in the original study) in cancer screening. Methods measuring tumour volume doubling time and using microsimulations were excluded. The references of the included studies were searched for relevant missing articles.

Because we aimed to identify estimates of overall mean sojourn time, for the updated search we reviewed all papers (included the ones in Geurts et al. (2022)) and further excluded those that did not report population-level estimates of (stage-)mean sojourn time, that reported estimates calculated from prevalence-incidence ratios only (further detail below), and that relied solely on empirical data that relates to screening tests with poor sensitivity for that cancer-site, including studies examining the use of X-rays for lung cancer screening.

We extracted information on the study, screening dataset, cancer type, mathematical model and sojourn time estimate(s). Mathematical models included: Maximum Likelihood Estimation (MLE), where a likelihood function built around a specific disease natural history models is maximised; Expectation-Maximization Algorithm (EMA), an iterative optimisation variation of MLE, used to handle latent data, in this case, the time a cancer enters its preclinical phase; Regression of Observed on Expected (ROE), an alternative regression-based technique that minimises the discrepancy between empirical screening observations and the expected outcomes of a natural history of disease model defined on sojourn time; and Bayesian models, which also define a natural history of disease model but use a distinct inferential approach that structurally integrates the primary data with prior probability distributions, allowing uncertainty to be integrated more seamlessly than other methods. We excluded prevalence-incidence ratio calculations, which arithmetically derives sojourn times strictly based on the ratio of preclinical cancer prevalence to clinical incidence. This method relies on stronger underlying assumptions, including that the screening test are 100% sensitive, and is known to be less accurate.

One researcher (DJ) conducted the updated literature search, selected and extracted the studies, and another (MS) confirmed the selection and extraction.

We summarised all estimates of interest (point estimate and associated uncertainty, where available) in each study, grouped by dataset and modelling approaches used to derive them (MLE, EMA, ROE or Bayesian).

**S1.2 Results**

From updating the review by Geurts et al. (2022), we identified and screened 332 unique records, identifying 9 new publications with estimates of mean overall sojourn time (Aarts, 2019; Bhatt, 2024; Choi, 2023; Weedon-Fekjaer, 2021; Wu, 2022; Ishizawa, 2024; Bhatt, 2021; Gonzalez Maldonado, 2021; Li, 2022) -- Table S1.2 and S1.3. Two additional studies were included, identified from content experts (Chen, 2001; Ryser, 2025). Combined with the original review, of which we used 15 studies (Walter, 1983; Day, 1984; Paci, 1991; Etzioni, 1997; Shen, 1999; Wu, 2005; Cong, 2005; Shen, 2005; Jiang, 2016; Shen, 2019; Alexander, 1989; Duffy, 1995; Myles, 2003; Launoy, 1997; Pinsky, 2001), we included a total of 26 studies across six cancer-sites; however, two of the studies in the original review report the same analyses and were considered together as the same study retrieving a final set of 25 studies (Walter, 1983; Day, 1984). Table 2 shows there is significant variation in the level of evidence across cancer-sites, with the most evidence existing for breast cancer.

Literature review: PRISMA diagram

**Figure S1.1.** PRISMA 2020 flow diagram for the updated literature review.

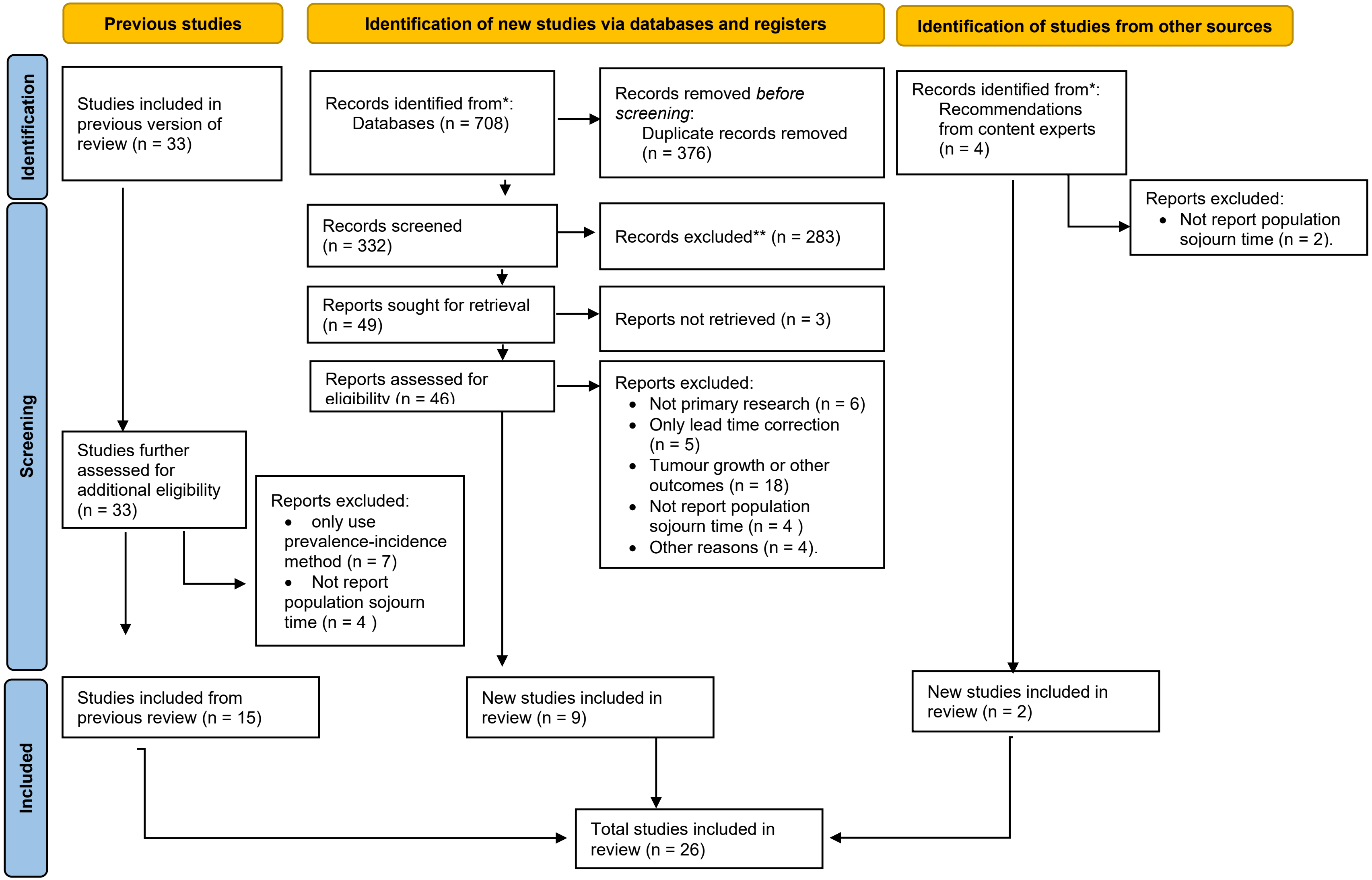

Included studies

A summary of the included studies that inform estimates of overall sojourn time can be found below. The 26 studies contribute with 52 estimates that vary in cancer type, dataset used, method of estimation and can also arise from the application of models under different assumptions.

**Table S1.2.** Studies included in the updated literature review.

| **Article (Author, years)** | **Source** | **Cancer site** | **Data source** | **Approach to sojourn time estimation** | **N overall mean sojourn time estimates** |
|---|---|---|---|---|---|
| Walter, 1983 and Day, 1984 | Original review | breast | HIP trial | MLE | 1 |
| Paci, 1991 | Original review | breast | HIP trial | ROE | 1 |
| Etzioni, 1997 | Original review | breast | HIP trial | EMA | 1 |
| Shen, 1999 | Original review | breast | HIP trial | MLE, | 1 |
| Wu, 2005 | Original review | breast | HIP trial | MLE; Bayesian | 2 (2 methods) |
| Cong, 2005 | Original review | breast | HIP trial; CNBS studies | MLE | 2 (2 datasets) |
| Shen, 2005 | Original review | breast | HIP trial | MLE | 1 |
| Jiang, 2016 | Original review | breast | HIP trial | MLE | 1 |
| Shen, 2019 | Original review | breast | HIP trial; Two-county trial | MLE (3 estimates from models with different assumptions) | 6 (2 datasets x 3 models) |
| Alexander, 1989 | Original review | breast | Edinburgh trial | MLE | 1 |
| Duffy, 1995 | Original review | breast | Two-county trial | ROE | 1 |
| Myles, 2003 | Original review | breast | Manchester (screened)+ Two-county trial (unscreened) | Bayesian | 1 |
| Launoy, 1997 | Original review | colorectal | Calvados study | MLE, Bayesian | 2 (2 methods) |
| Pinsky, 2001 | Original review | colorectal | Calvados study | MLE (2 estimates from models with different assumptions) | 2 (2 models) |
| Aarts, 2019 | Update | breast | Nijmegen 2007-2012; 2001-2006; 1989-2000; 1975-1988 | MLE; NLR; Bayesian | 12 (4 datasets x 3 methods) |
| Bhatt, 2024 | Update | breast | SEARCH trial | MLE | 1 |
| Choi, 2023 | Update | breast | Korean screening programme | MLE | 1 |
| Weedon-Fekjaer, 2021 | Update | breast | BreastScreen (Norway) | MLE | 1 |
| Wu, 2022 | Update | breast | HIP trial | MLE; Bayesian | 2 (2 methods) |
| Ishizawa, 2024 | Update | ovarian | PLCO trial, UKCTOCS trial; USS; MMS | Bayesian | 3 (3 datasets) |
| Bhatt, 2021 | Update | prostate | PLCO trial | MLE | 1 |
| Gonzalez Maldonado, 2021 | Update | lung | LUSI trial | MLE | 1 |
| Li, 2022 | Update | colorectal | FIT pilot, f-Hb 20, f-Hb 120, f-Hb 180 | MLE | 3 (3 datasets) |
| Chen, 2001 | Advice from experts | liver | Taiwan data, liver cirrhosis, non-cirrhosis | ROE | 1 |
| Ryser, 2025 | Advice from experts | ovarian | UKCTOCS trial/USS; MMS | Bayesian | 2 (2 datasets) |

* ROE: Regression of Observed on Expected; MLE: maximum likelihood estimation; Bayesian: Bayesian MCMC simulation; EMA: expectation-maximization algorithm; NLR: non-linear regression

#### S2. Elicitation protocol

#### Protocol for estimating stage-sojourn time of cancer: review and elicitation of expert opinion to inform natural history of disease models of Multi-Cancer Early Detection tests

*Dina Jankovic, Matthew E.J. Callister, Georgios Lyratzopoulos, Stephen Palmer, Marta O. Soares*

V1: 09.10.2025

The Galleri® (GRAIL) blood test is a multi-cancer early detection (MCED) test that detects circulating tumour DNA (ctDNA) signals shared by over 50 cancers (Liu et al., 2020). The test aims to improve outcomes by early detection of cancers, when treatment is likely to be most effective.

A large, randomised trial, the NHS-Galleri trial, is being conducted in the UK to examine whether the late-stage cancer detection rate can be reduced by adding a screening regimen with the Galleri test (three annual screens in asymptomatic population aged 50-77) to current care. The impact of a stage shift observed in the trial is likely to vary across cancer types. The policy decision on the wider use of the Galleri test will need to reflect which cancer types drive the observed stage-shift to determine expected value of the test. It will also need to examine different ways in which the Galleri test could be implemented in the NHS (e.g. different screening intervals, different risk groups, etc) to optimise stage-shift, value and efficiency. Alongside the NHS-Galleri trial, a policy decision will therefore need to consider the natural history of disease, and any differences across cancer types.

A mathematical model of the natural history of disease will reflect the duration of undetected but detectable cancer (UDC). UDC starts when a cancer becomes detectable as cancer (or screen-detectable) and lasts until a cancer is detected (symptomatic, incidental or via the routine screening programmes that are part of current care) or the individual dies from other causes. The definition of UDC has been chosen to represent the quantities that can be inferred from screening data –and therefore that will conform with the modelling of NHS-Galleri data required for the Galleri test evaluation. It is also consistent with what has been measured in previous studies that used primary screening data to estimate, what they refer to as cancer sojourn times. To maintain consistency with the existing literature, throughout the elicitation we use the term *sojourn time* to describe the UDCs but we are very clear about how we define it and the implications, including how these quantities are affected both by cancer characteristics and clinical practice.

The extent of a stage-shift, and the value of a screening test, can depend on the cancer stage-sojourn time, i.e. on the duration of time cancers spend in each preclinical stage before progressing to a more advanced preclinical stage or being detected. Cancers with lower stage-sojourn times (relative to the screening interval) are more likely to be missed by screening, but when identified early may show largest stage shifts. In this study, we will consider two stage-sojourn times: early-stage (usually stage I and stage II) mean sojourn time (EMST) and late-stage (usually stage III and stage IV) mean sojourn time (LMST). Overall mean sojourn time (OMST) is the sum of early and late-stage sojourn times, while accounting for the fact that some cancers never enter late stage.

MCED tests can identify more than one cancer type and, due to the heterogeneity in stage-sojourn time, their use for screening is likely to lead to different stage-shifts, and produce different value, across different cancer types. The ongoing large clinical trial, the NHS-Galleri trial, has been powered on an aggregated estimate of stage-shift (reduction in late-stage cancer incidence) across multiple individual cancer types, and so evidence from the trial is unlikely to be sufficient to inform natural history modelling for individual cancer types.

Stage-sojourn time (as per the definition used here) is inherently unobservable and not directly estimable. It can, however, be inferred from screening data, or predicted from examining the evolution of cancer characteristics over time such as tumour growth rates. We reviewed existing MCED natural history of disease models (Hubbell et al., 2021; Tafazzoli et al., 2022; Sasieni et al., 2023; Dai et al., 2004, Lange et al., 2024) and identified that these use expert opinion and estimates from the literature to inform stage-sojourn times. The estimates informed by experts (Hubbell et al., 2021; Tafazzoli et al., 2022; Sasieni et al., 2023; Dai et al., 2004) were elicited in a study where sojourn times were not the primary focus and so the process was not optimised. For example, the definition of sojourn times was not clear, experts were provided limited background information on which to base their estimates, the estimates did not capture experts' uncertainty and the sample only included US-based experts. MCED models that used literature-derived estimates of sojourn times (Dai et al., 2004, Lange et al., 2024) use selected (single) estimates with limited justification, potentially missing relevant available literature.

Furthermore, none of the existing MCED models reflect complexities such as correlation in sojourn times across different stages, and none of the estimates are specific to ctDNA cancers detected by the Galleri test, considered to be potentially more progressive and therefore likely to have shorter sojourn times. Overestimating the sojourn time has the potential to overestimate stage shift with screening.

In this work we propose to use formal structured expert elicitation (SEE) supported by best available evidence to characterise expected sojourn times, together with uncertainty estimates, across cancer types in the UK setting. We will elicit sojourn times for all cancers (ctDNA positive and negative) and for ctDNA-positive cancers, separately. The work will address limitations of previous sojourn time estimates used in existing MCED models by quantifying uncertainty around the estimates, by taking into account complexities such as stage sojourn time being conditional on cancer stage at diagnosis, and by using a range of relevant evidence to support the elicitation process. The elicited estimates will be used to provide informative prior distributions for natural history modelling using data from the NHS-Galleri trial.

The methods in this protocol were developed in line with reference methods for SEE in HTA (Bojke et al., 2022).

**What do we want to elicit?**

We aim to elicit, with uncertainty, OMST by cancer type for all cancers (ctDNA positive and negative combined) and for ctDNA cancers, and stage- mean sojourn time (stage-MST) by cancer type for all cancers. We define OMST as the average duration of time cancers are undetected (at any stage) with current care, including symptom-led and incidental diagnoses, and detection via existing screening programmes as these are part of usual care, but should not include diagnoses from the Galleri test.

Stage-MST is the average duration of time cancers spend in each preclinical stage before progressing to a more advanced preclinical stage or being clinically detected. We will elicit two cancer stage-MSTs – EMST and LMST.

We also aim to examine the level of variation in overall sojourn time across individuals (which is often termed heterogeneity in this literature). This will help validate whether overall sojourn time follows an exponential distribution (i.e. constant risk of progression over time), typically assumed in empiric estimates of sojourn time.

**What evidence will be provided to experts, to support the elicitation (evidence dossier)?**

Before answering any questions, experts will be provided with definitions of sojourn times and a summary of evidence sources and evidence gaps.

Critically, to ground the elicitation, experts will also be provided with the following evidence, for each cancer type:

- empiric estimates of overall mean sojourn times informed by a review conducted for this purpose (further detail below);

- five-year survival and staging at diagnosis of clinically detected cancers and how it compares to other cancer types from National Cancer Registration and Analysis Service (NCRAS) (NCRAS 2025a, NCRAS 2025b);
- screening test, screening interval and target population of cancer screening programmes (breast, colorectal, lung and cervical);
- test sensitivity by cancer type and stage (in early stage, late stage and overall), estimated from the Galleri test sensitivity in the CCGA3 study (Klein et al., 2021), to provide an indication of the proportion of cancers that are ctDNA,
- a summary of evidence on ctDNA cancers, including the following:
    - Overall cancer survival for cancers detected vs not detected by the MCED, compared to SEER expected survival adjusted for cancer type, stage, age and gender. The survival estimates will be stratified by cancer stage or by cancer type (for breast, colorectal, lung and prostate cancers) (Chen et al., 2021; Swanton et al., 2025).
    - ctDNA detection rate 1, 2 and 3 years prior to conventional cancer diagnosis, based on samples donated by healthy study participants, at baseline (Patel et al., 2023a; Patel et al., 2023b)

Empirical estimates of OMST were derived from a literature review of mathematical estimates derived from the analyses of primary data either from clinical trials of screening interventions or from routine data from existing screening programmes (Geurts et al., 2022) that we have updated to February 2025. Using search terms and inclusion criteria from the original review, we identified and screened 332 unique records, identifying 13 new papers on OMST. Combined with the original review, we had 43 studies for 6 cancer types, summarised in Table 1. This is here considered the most robust evidence of OMST available and, to ground the elicitation, we will summarise point estimates and uncertainty from each study, grouped by datasets and modelling approaches used to derive them.

Table 1. Number of studies identified in the literature review, by cancer type.

| Cancer type | Original review | Update | Total |
|---|---|---|---|
| Breast | 22 | 7 | 29 |
| Lung | 2 | 2 | 4 |
| Colorectal | 2 | 2 | 4 |
| Cervical | 4 | 0 | 4 |
| Ovarian | 0 | 1 | 1 |
| Prostate | 0 | 1 | 1 |
| Total | 30 | 13 | 43 |

**What quantities to elicit?**

A key consideration in defining the specific quantities to be elicited is to ensure expert workload is manageable. Given the multicancer nature of the technology, which includes 21 stageable cancer types (as considered in previous research, Sasieni et al., 2023), we adopted a number of simplifications to reduce the number of cancer-type specific estimates elicited.

Firstly, we prioritised quantitative elicitation for the subgroup of 11 cancer types in which screening with the Galleri test is expected to produce the highest health gains (colon and rectum, lung and bronchus, ovarian, head and neck, lymphoma, pancreas, prostate, breast, liver and intrahepatic bile duct and oesophagus, (Kunst et al., under review) or for which empirical estimates of sojourn times are available (Table 1). For the remaining 10 cancers, sojourn times will be elicited qualitatively, by asking experts to select one or more of the previously elicited cancer types they are most similar to.

Secondly, stage-MST will be elicited for the six cancers for which empirical estimates of sojourn times are available. For the remaining 15 cancers, assumptions will be made about the duration of stage-MSTs relative to OMST.

Finally, uncertainty, variability and ct-DNA specific estimates will be elicited for OMST only. The assumptions used to extrapolate these to stage-MST are described below.

The quantities to be elicited are summarised in Table 2.

Table 2. Quantities to be elicited.

| Question number | Quantity | Rationale |
|---|---|---|
| Q1a | OMST for all cancers combined (ctDNA positive and negative) (elicited with uncertainty) | The only sojourn time parameter with empirical estimates. SEE will capture experts' beliefs about the validity and generalisability of the empirical estimates. |
| Q1b | OMST for cancer types with a screening programme (breast, colorectal, cervical and lung), including screen-detection (elicited with uncertainty) | Informs OMST of all cancers in the UK setting, in absence of the Galleri test. Unlike Q1a, this estimate includes the sojourn time of screen-detected cancers. |
| Q1c | 25th percentile of overall sojourn time (point estimate only)<br><br>This is elicited conditioned on assuming the mode of Q1a is the true OMST | - To be used to describe heterogeneity in overall sojourn time and validate exponential or fit alternative distribution.<br>- Fitted distribution to be used to estimate MST for the x% of the cancers with the shortest sojourn time.* |
| Q2a | OMST of cancers diagnosed in early stage (point estimate only)<br><br>This is elicited conditioned on assuming the mode of Q1a is the true value | - Informs (equivalent to) EMST of cancers diagnosed in early stage.<br>- Can infer OMST of cancers diagnosed in late stage using the % diagnosed in late stage and the OMST from Q1a. |
| Q2b | EMST for cancers diagnosed in late stage (point estimate only)<br><br>This is elicited conditional on OMST for cancers diagnosed in late stage inferred in Q2a | Informs EMST and LMST (OMST - EMST) for cancers diagnosed in late stage. |
| Q3 | OMST for ctDNA cancers (elicited with uncertainty)<br><br>This is elicited conditional on assuming that the mode of Q1a is the true value. | Qualitative questions to establish whether Galleri detected cancers are those with shortest sojourn time and, if so, another qualitative question validating prediction using estimates from Q1c. If none of the previous, elicit with uncertainty. |

* x represents the proportion of cancers that are ctDNA positive, estimated from the Galleri test sensitivity in the CCGA3 study (Klein et al., 2021)

To estimate uncertainty in EMST and LMST, we will propagate the elicited uncertainty in OMST. EMST and LMST in ctDNA cancers will be derived from answers to Q1a, Q2a and Q2b, assuming equal ratios between EMST, LMST and OMST.

At the end of each question we will ask experts to provide the rationale for their answers and additional comments about the quantities (e.g. caveats to their answers) in free text. At the end of the exercise, we will ask experts to comment on the clarity of the questions, to state if it was difficult or challenging to reply to the questions, and we will allow them to express any additional comments in free text.

**Which experts?**

We propose to elicit from clinical experts acting as substantive experts in the disease area. We will explore, in a pilot, the suitability of recruiting medical professionals from different areas of specialism. While specialists may be able to provide more precise estimates of sojourn times in cancer types within their specialty. Generalists may have a better understanding of relative MSTs between different cancer types. Experts will be purposely selected, e.g. through publications (relating to sojourn times or ctDNA cancers), participation on relevant advisory boards (e.g. the Galleri trial, the National Screening Committee) and on recommendation by peers.

**How to elicit?**

Method of elicitation

For quantitative elicitation with uncertainty, we will use the chips and bins method (O'Hagan et al., 2006) where experts are provided with a grid that divides the expert's plausible range into intervals and are asked to construct a histogram, with each 'chip' representing a unit of probability (e.g. 20 chips each worth 5%) placed into one of the intervals or 'bins'. It is considered suitable for clinical experts, and more intuitive than other available methods (Bojke et al., 2021). The method was chosen after all three experts in the pilot found it more engaging and preferable than the direct elicitation of 80% confidence intervals.

Quantitative elicitation without uncertainty will request individuals to indicate the value they believe is most plausible (point estimate, mode). In the analysis, we will explore the impact of uncertainty through scenario analyses.

The wording of the questions will be piloted for clarity and adequacy. The time it takes to complete the exercise will be recorded in the pilot.

Model of delivery

The elicitation will be conducted in remote, live video conference sessions lasting up to two hours. In the sessions, experts will be provided training, relevant background evidence and an opportunity for discussion. The experts will answer the elicitation questions within the live sessions, individually, with an opportunity to complete any unanswered questions independently.

We will use a bespoke online tool to collect experts' responses. This will allow us to condition the formulation of a particular question to the responses to previous questions and to ensure we can highlight any inconsistencies (e.g. if an expert enters mathematically incoherent values).

Training of experts

Training provided during the live session will have the following aims: describe the objectives of the elicitation exercise, clarify concepts such as those of uncertainty, variability and heterogeneity, familiarise the experts with the quantities we wish to elicit, describe and explain the impact of bias and heuristics with a special focus on overconfidence, and train experts on the method of elicitation used.

**How are the judgements provided processed and analysed?**

Approach to pooling judgements from multiple experts

When several experts supply probability distributions, their responses are often pooled to derive a single distribution. We will use a mathematical approach where we analytically pool the individual answers. This approach is used instead of a consensus approach because consensus methods are known to have a number of limitations (e.g. implicit aggregation, effect of dominant individual on group dynamics, known problems where consensus returns overly-precise judgements) (Bojke et al., 2022). We will linearly pool across experts using equal weights. Linear pooling preserves the individual judgements in the collective (pooled) judgement. For example, if the experts' distributions for a single quantity are identical the pooled is equal to the individuals' distributions. Also, if there is the support from at least one expert that the quantities of interest take particular values, the pooled distribution will also show some support for those values (O'Hagan et al., 2006).

Fitting of distributions

We will fit a distribution to the elicited summaries to represent uncertainty. We will fit a range of distributions (e.g. Exponential, Weibull, Lognormal and Gamma) to the overall mean sojourn time to constrain the range to non-negative values and select the distributions with the best fit for the analysis.

##### Pilot

The materials were first reviewed by one non-substantive expert clinician (general practitioner) and a pilot was run with two substantive expert clinicians (a specialist oncologist, MC, and a cancer epidemiologist specialising in early diagnosis research, GL). Following feedback from the pilot with the expert clinicians, the protocol was updated to specify the method for eliciting uncertainty, clarify the definition of cancer sojourn times, to add detail to graphs showing published estimates of OMST and to elaborate on evidence regarding prognosis and detectability of ctDNA cancers.

##### Data protection and anonymity

Experts will be asked to give their opinions individually (not in groups). The information provided will be pseudonymised, stored securely and only accessed by those carrying out the study.

#### S3. Training slides

### Structured expert elicitation of cancer sojourn times

7th October 2025

#### Overview

| Topic | Slide No. |
|---|---|
| Background – 20min | 3 |
| Training and practice – 10min | 29 |
| Questions – 90min | web app |

#### Overview

| Topic | Slide No. |
|---|---|
| Background – 20min | 3 |
| Training and practice – 10min | 29 |
| Questions – 90min | web app |

#### Project background

**Project aim:** to evaluate the clinical and cost-effectiveness of the Galleri® (GRAIL) blood test, a multi-cancer early detection (MCED) test that detects circulating tumour DNA (ctDNA) signals shared by over 50 cancer types.

**Why do we need information on sojourn times**: To model the natural history of disease across cancer types.

- The NHS-Galleri trial is powered on 'all cancer' outcomes, and cannot distinguish cancer types.
- Understanding the value of an MCED test requires consideration for **how cancers differ in sojourn time**, or the duration of time they spend undiagnosed

#### Terminology

the quantities we are interested in eliciting

##### Duration of undetected but detectable cancer

This is the duration of time cancers spend in the detectable phase, before being diagnosed :

- Detectable phase starts when a cancer can be detected as cancer,
- Diagnosis can refer to symptomatic, incidental or screen-detection.

For simplicity, we refer to this quantity throughout as **sojourn time**.

### Terminology

diagnosis, symptomatic detection

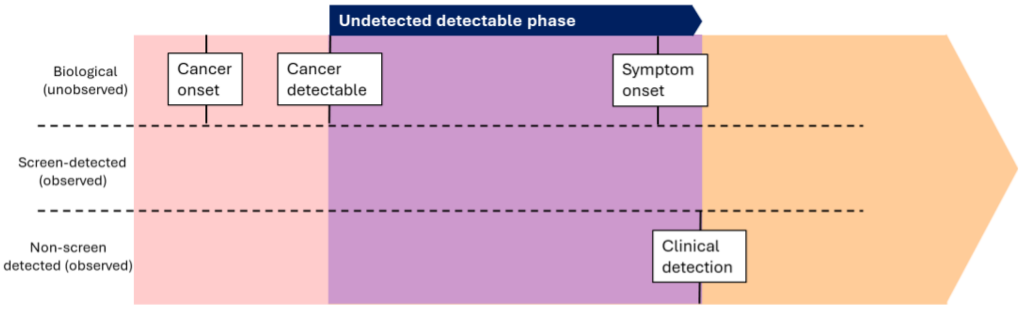


### Terminology

diagnosis, incidental detection

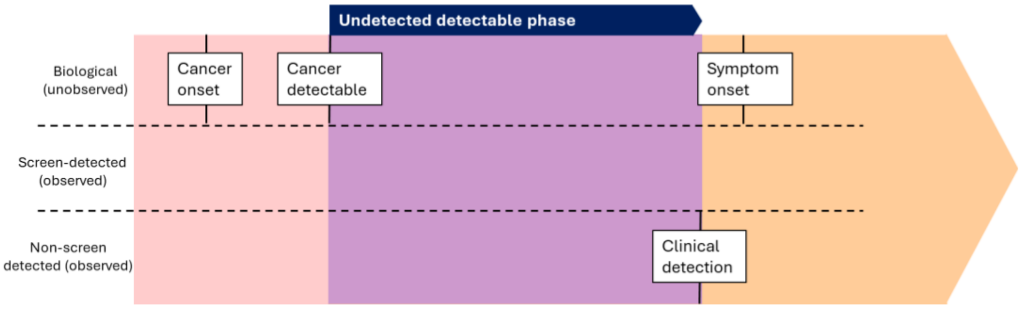

### Terminology

diagnosis, screen-detection

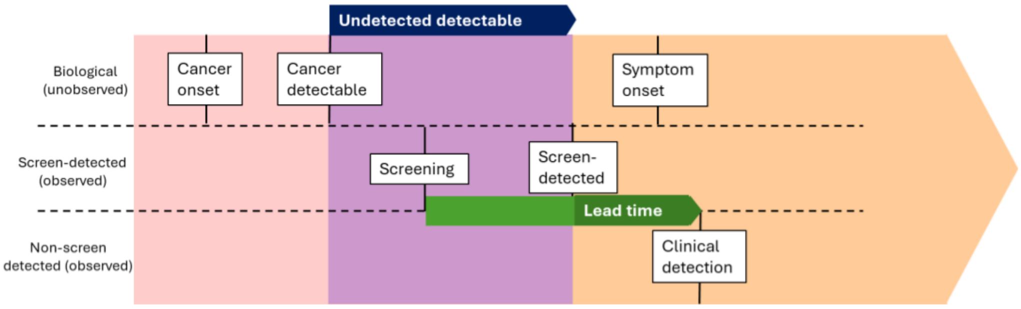


Only applicable to cancers with existing screening programmes (colon/rectum, breast, cervix).

### Terminology

specific quantities to elicit

- **Overall mean sojourn time (OMST):** mean (across all individuals that develop cancer) of the total duration of undetected but detectable cancer (i.e. across all stages up to detection)

- **Stage-sojourn times:** duration of time cancers spend in different stages before they progress to a later stage, are detected. In this study, we will consider two stages – early (usually stages I and II) and late (usually stages III and IV)
  - Early-stage mean sojourn time (**EMST**)
  - Late-stage mean sojourn time (**LMST**)

### Terminology

OMST and stage-sojourn times

… for cancers identified in early stage, OMST = EMST

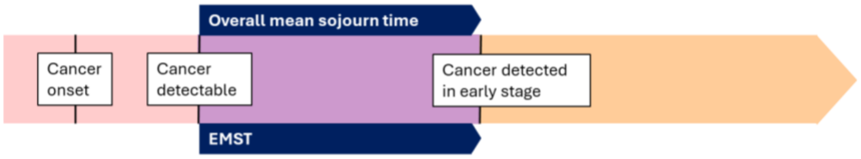


… for cancers identified in late stage, OMST = EMST + LMST

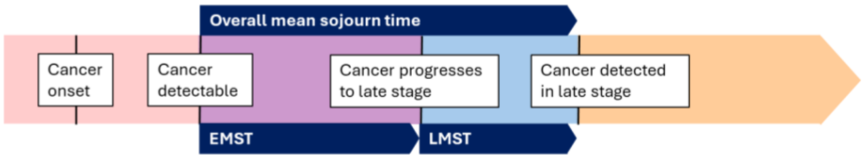

### Terminology

What determines sojourn times?

**Cancer characteristics such as …**

- Cancer aggressiveness: more aggressive cancers ↓sojourn times
- Prominence of symptoms: e.g. ability to identify breast nodules from palpation means shorter sojourn time than, for example, lung cancer

**Clinical practice, for example…**

- Likelihood of detection at earlier stages: e.g. better resolution of available tests prolongs detectable phase, leading to longer sojourn times.
- Public awareness of symptoms: e.g. increase in awareness of breast cancer symptoms may have decreased time to diagnosis, leading to shorter sojourn times
- Availability of cancer screening programmes: cancers with screening programmes as part of routine care are intercepted early, leading to shorter sojourn times
- Health system factors such as limitations in access to services or large waiting times can affect time to detection and therefore sojourn times

### Empiric estimates of sojourn times

1. NHS-Galleri trial
   - powered to detect aggregate outcomes across multiple individual cancer types, unlikely to inform sojourn time for individual cancer types.
2. Existing MCED models
   - elicited estimates: process not optimized to eliciting sojourn times
   - literature-based estimates use selected (single) estimates with limited justification

2. Empiric evidence

#### Empirical estimates of sojourn times
preferred estimation approach

Inference from screening data, mathematical estimates

- Primary data from screening trials or routine data on screening programmes
- Data on detection at multiple screening rounds and detection between screening rounds and/or in the absence of screening used to mathematically estimate sojourn times
- Account for complexities such as imperfect screening test sensitivity

#### Empirical estimates of sojourn times
other evidence

e.g. inference from calibration models, where it is less clear how sojourn times are determined/evidenced

e.g. predictions based on the evolution of untreated cancer characteristics over time, e.g. tumour growth rate

#### Empirical estimates of sojourn times

##### review of mathematical estimates

We updated a published literature review (Geurts et al., 2022) to 2025

| Cancer type | Original review | Update | Total |
|---|---|---|---|
| Breast | 12 | 7 | 19 |
| Lung | 0 | 1 | 1 |
| Colon/Rectum | 2 | 1 | 3 |
| Ovary | 0 | 2 | 2 |
| Prostate | 0 | 1 | 1 |
| Liver | 0 | 1 | 1 |
| Total | 14 | 13 | 27 |

#### Empirical estimates of sojourn times

##### evidence gaps

- OSMT estimates exist for **six** cancer types (breast, lung, colon/rectum, ovary, prostate and liver)
- Variation in OMST estimates between studies, unclear which is most relevant to the UK setting
  - Studies use different datasets and estimation methods
- No estimates specific to ctDNA cancers
- Stage-sojourn times estimates exist for one cancer type (ovary)

#### Structured expert elicitation
aim of today's session…

… to use **structured expert elicitation (SEE)**, supported by best available evidence, to characterise mean sojourn times, and uncertainty over these, across cancer types in England.

#### Structured expert elicitation
what is SEE?

***"systematic process of formalizing and quantifying, typically in probabilistic terms, expert judgments about uncertain quantities"***

Uncertainty arises from….

- Evidence being sparse or not entirely generalizable,
- Observation/experience being limited or less relevant,
- Limited mechanistic knowledge.

Uncertainty is not bad,

- We make 'better' decisions if we quantify this uncertainty and incorporate it into our decision-making processes

Structured methods minimise bias, improve transparency and accountability.

#### **Structured expert elicitation**
#### what will we ask you to do?

- We will provide you with some training on how to express your uncertainty
- We will present evidence base that we identified as relevant for estimating cancer sojourn times.
- We will ask you a series of questions relating to cancer sojourn times
  - For some quantities we will ask you about your uncertainty
- Should take up to 2 hours.
- All responses will be anonymised.

#### **Structured expert elicitation**
#### different information elicited across cancer types

| **Questions** | **Cancer types** |
|---|---|
| OMST and stage-OSMT for all cancers, OMST for ctDNA cancers | Six cancer types for which empiric estimates of OMST are available: breast, lung, colon/rectum, prostate, ovary, liver/bile duct |
| OMST for all and ctDNA cancers | Five further cancer types in which screening with the Galleri test is expected to produce the highest health gains: cervix, head and neck, lymphoma, oesophagus, pancreas |
| Multiple choice questions about OMST | Remaining ten cancer types: anus, bladder, gallbladder, kidney, melanoma, sarcoma, stomach, thyroid, urothelial tract, uterus |

**Structured expert elicitation**
different information elicited across cancer types

- OMST for “all” cancers
- Within cancer type variation in sojourn time for “all” cancers
- stage-OSMT for “all” cancers
- OMST for ctDNA cancers

We will ask about all cancer types for which you can provide insight.

Overview

| Topic | Slide No. |
|---|---|
| Background – 20min | 3 |
| Training and practice – 10min | 28 |
| Questions – 90min | web app |

#### Expressing uncertainty

How will I be asked to express uncertainty?

- For some questions we will ask you to express your uncertainty
- We use the Chips and Bins method
- Practice question: what proportion of UK households own a pet?

#### Chips and Bins: first elicit the plausible limits

| | |
|---|---|
| LOWER PLAUSIBLE LIMIT (L) | A value such that you believe that there is a **1% probability** that the true value is less than L |
| UPPER PLAUSIBLE LIMIT (U) | A value such that you believe that there is a **1% probability** that the true value is greater than U |

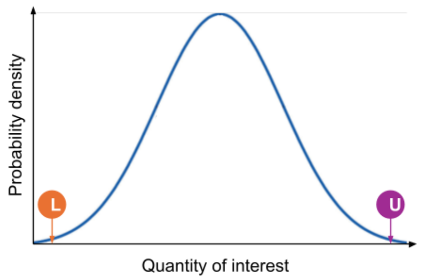


**Why start with plausible limits?**

- Minimizes the bias of **over-confidence** – not assigning enough probability to extreme values
- Minimizes the bias of **anchoring** – assigned too much credibility to a value already in the expert's mind
- Provides **bounds** to the uncertainty

25

### Chips and Bins

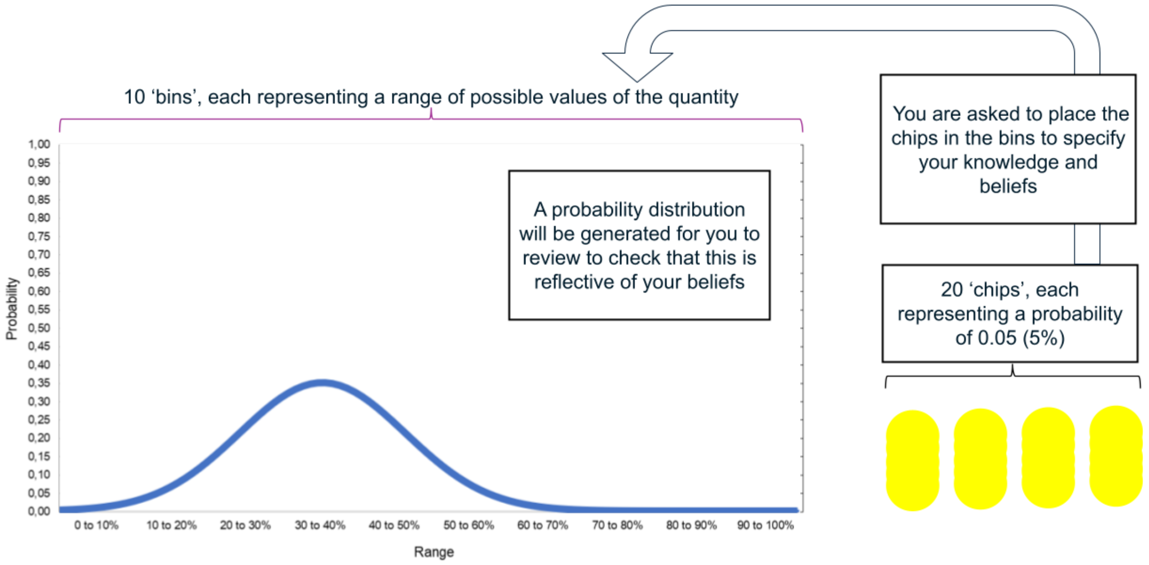


26

### Chips and bins: Example 1.

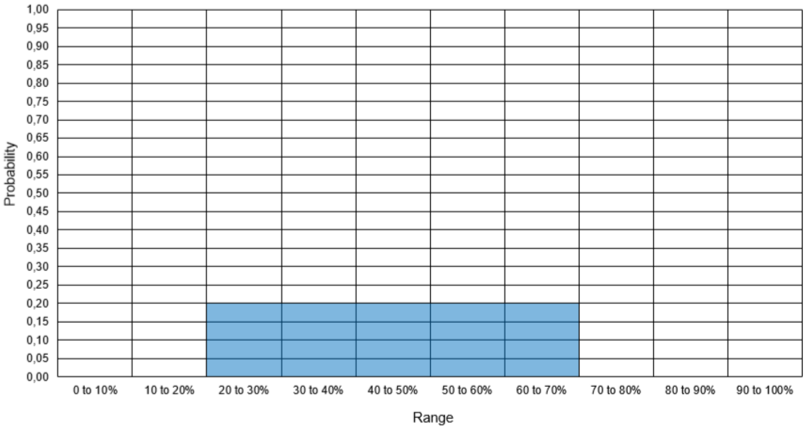


**Example 1**
You are certain that the true value lies between 20% and 70% but it is equally likely to be any of the values in between

27

### Chips and bins: Example 2.

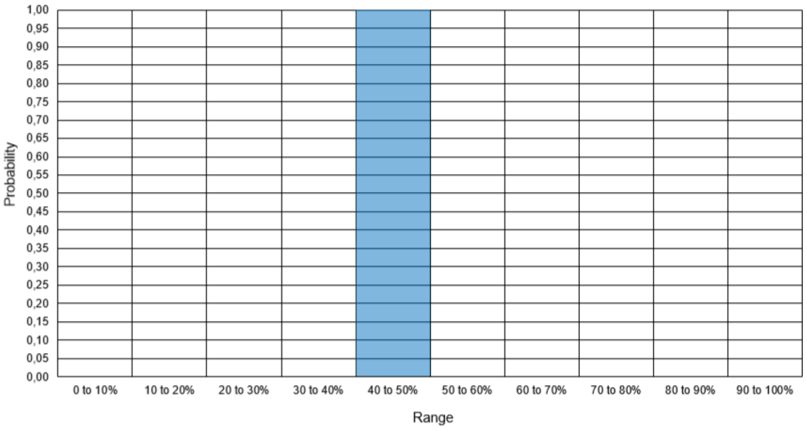


**Example 2**
You are certain that the true value lies between 40% and 50%. No other values are possible.

28

### Chips and bins: Example 3.

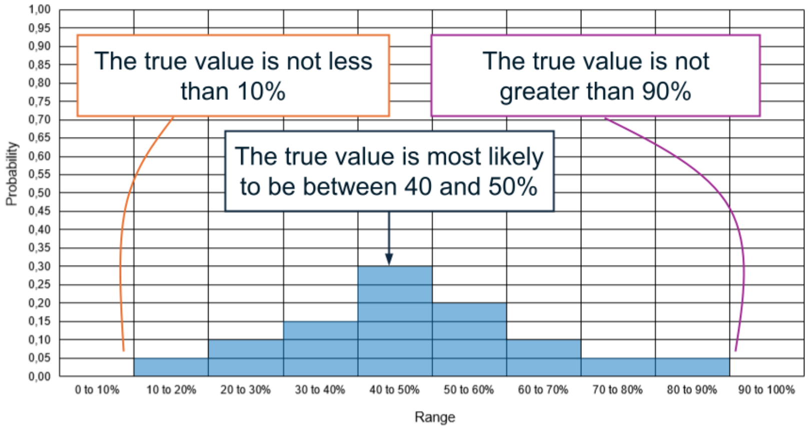


**Example 3**
The true value is equally likely to fall within the following ranges:

- <40%
- 40–50%
- 50–70%

There is a smaller chance that the true value is >70%

29

#### Practice questions

Placeholder: please go to https://shiny.york.ac.uk/cancer_sojourn_time/ to complete your practice question.

You will then go straight to the elicitation questions in the app.

#### Overview

| Topic | Slide No. |
|---|---|
| Background – 20min | 3 |
| Training and practice – 10min | 28 |
| Questions – 90min | web app |

#### S4. Elicitation questions (including relevant background information): Breast cancer example

##### Breast cancer prognosis in England

| **Five-year survival (bar, left-side axis) and number of cases between 2015-2019 (line, right-side axis).** *Cancer types ordered by largest to smallest survival* | **Breakdown of early vs. late stage at diagnosis** *Cancer types ordered from largest to smallest proportion of early-stage cancers at diagnosis* |
|---|---|
| 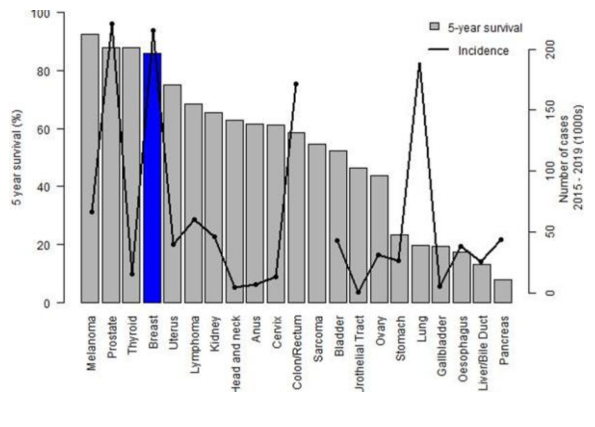 | 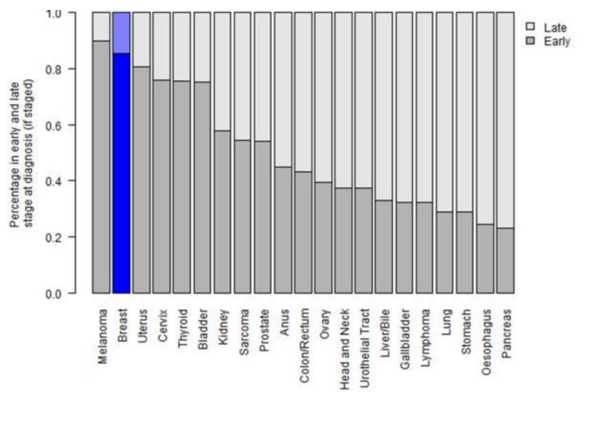 |





##### OMSTs reported in the literature

*Studies identified in a literature review of mathematical estimates derived from the analyses of primary data either from clinical trials of screening interventions or from routine data from existing screening programmes (Geurts et al., 2022) that we have updated to 2025.*

#### Summary of OMST estimates

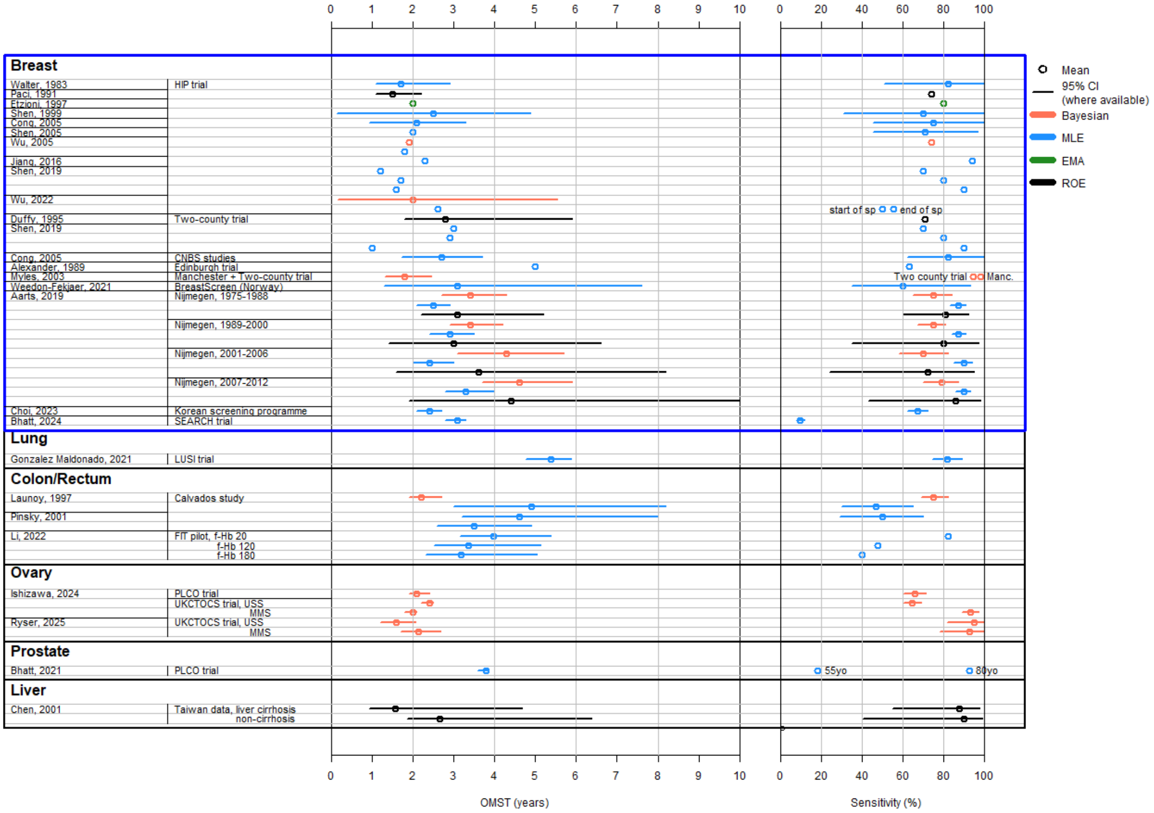

#### Summary of datasets

| Cancer type | Dataset | Country | Age | Time period | Screening strategy |
|---|---|---|---|---|---|
| Breast | The Health Insurance Plan of Greater New York (HIP) randomized trial | US | 40-64 | 1963-1966 | Three annual mammograms (analogue) + clinical examination. One year follow up. |
| | Two-County trial | Sweden | 40-74 | 1977-1984 | 40-49 years: biennial mammogram, for up to 7 years.<br>50-74 years: mammogram every two to three years, for up to 7 years.<br>Single view mammogram later changed to two-view mammogram.<br>Recruited individuals with family history of breast cancer. |
| | Edinburgh Randomized Trial of Breast Cancer Screening | UK | 45-64 | 1978-1988 | One two-view mammogram + clinical examination, followed by six more annual clinical examinations |
| | Nijmegen breast screening programme data | The Netherlands | 50-69 | 1975-2012 | Biennial mammogram, details changed over time<br>- 1975 – 1988 analogue mammogram, within trial;<br>- 1989 – 2000 analogue mammogram;<br>- 2001 – 2006 analogue mammogram, increased sensitivity;<br>- 2007 – 2012 digital mammogram |
| | SEARCH study participants + NHS Breast Screening Programme records | UK | 47-70 | 1989-2019 | Mammogram every three years as per UK screening programme (technique changed over time) |
| | Korean National Cancer Screening Program data from tertiary hospitals | South Korea | 40-70 | 2009-2013 | Biennial mammogram. |
| | BreastScreen Norway data | Norway | 50–69 | 1995-2002 | Biennial two-view mammogram. |
| | Manchester screening programme study | UK | 19-71 | 1987-1997 | Annual mammogram, superceded by bianneal mammograms in later years, offered to individuals with family history of breast cancer. |
| | Canadian National Breast Screening Studies | Canada | 40-59 | 1980-1985 enrollment (+ mean follow up 8.3 years) | Annual mammogram and physical examination |
| Lung | LUSI trial | Germany | 50-72 (male smokers) | 2007 - 2015 | Five rounds of annual low dose computer tomography (LDCT) |
| Colon/Rectum | FIT pilot study | UK | 59–75 | 2014 | Bianneal Faecal Immunochemical Test (FIT). Explored different thresholds for faecal haemoglobin concentration (f-Hb) but 120 µg/g threshold rolled out. |
| | Calvados study | France | 45-74 | 1991-1994 | Fecal occult blood test every two years. |
| Ovary | PLCO trial, SEER data | US | 55 - 74 | 1993-2001 enrollment (+ max 13 years follow up) | Annual CA-125 blood test for six years + annual transvaginal ultrasound for four years (performed independently) |
| | UKCTOCS trial, SEER data | UK | 50–74 | 2001-2005 enrollment | Multimodal screening: annual serial CA-125 measurements + second-line transvaginal ultrasound if high risk (max 11 rounds)<br>Ultrasound screening: annual transvaginal ultrasound (max 11 rounds) |
| Prostate | PLCO trial, SEER data | US | 55 - 74 | 1993-2001 enrollment (+ max 13 years follow up) | Six annual Prostate-Specific Antigen tests + four annual Digital Rectal Examinations |
| Liver | Taiwan 2-stage screening programme data | Taiwan | NR (55% < 50 years old) | 1991-1998 | Abdominal ultrasound screening offered to individuals with at least one of the following six : hepatitis B surface antigen, antibody for hepatitis C , alpha-feroprotein (AFP)≥ 20ng/mL, aspartate transaminase ≥ 40 IU/L, alanine transaminase ≥ 45 IU/L and family history of hepatocellular carcinoma.<br><br>Screening interval:<br>- three months if diagnosed with hemangioma, pseudotumor, AFP ≥ 20 ng/mL and liver cirrhosis;<br>- six months if early liver cirrhosis;<br>- one year in none of the above. |

**Question 1: OMST in all cancers (ctDNA and non-ctDNA)**

**Question 1a: What is the overall mean sojourn time (OMST) of clinically detected breast cancers in England?** (elicited with uncertainty using Chips and Bins)

**Question 1b: What is the overall mean sojourn time (OMST) of breast cancers in England?** *This includes clinically and screen-detected cancers.* (elicited with uncertainty using Chips and Bins)

**Question 1c:**

In this question we are interested in how much sojourn times vary between cancers of the same type.

**I believe that, if mean sojourn time of breast cancer is << *INSERT MODE from Q1b*>> years, then 25% of patients with the fastest-progressing cancers (shortest sojourn time) will have a sojourn time of ______ years or shorter.**

**Question 2: Mean stage sojourn times in all cancers**

**Q2a: What is the OMST for cancers diagnosed in early stages?** *Note that this is the same as early-stage mean sojourn time (EMST) for cancers diagnosed in early stages.*

If OMST for breast cancer is << *INSERT MODE from Q1b*>> years, and 15 % of cancers will progress to late stage before being diagnosed, what is the OMST in cancers diagnosed in early stage?

Feedback provided: Your answers imply that OMST for cancers diagnosed in late stages is <<DERIVE from Q1b and Q2a>> years.

**Q2b: What is the EMST for cancers diagnosed in late stages?**

If cancers diagnosed in late stages have an overall sojourn time of <<DERIVED VALUE from Q1b and Q2a>> years, how much of that time will those cancers have spent in early-stage?

Feedback provided: Your answers imply that late-stage mean sojourn time is <<DERIVED VALUE from Q2a and Q2b>> years.

#### Question 3: OMST in ctDNA cancers

##### Survival for cancers detected vs not detected by Galleri

- Participants who developed cancer in the first 3 years of follow-up in CCGA study (Chen et al., 2021; Swanton et al., 2025).
- Participants' blood samples were tested for cancer signals post-diagnosis.
- Outcome: overall survival for cancers detected vs not detected by Galleri (stratified by stage), compared to SEER expected survival adjusted for cancer type, stage and gender mix.
- Note: the values include all cancer types.

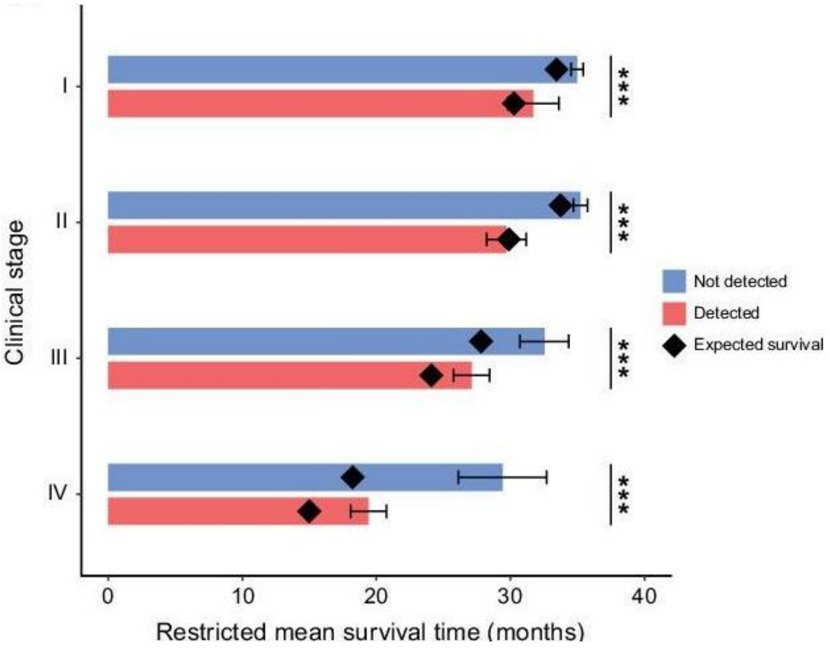


##### Signal detection in samples prior to cancer diagnosis

- Participants who enrolled in the study healthy and donated blood samples at the start (these were frozen).
- Samples analysed for participants who developed cancer within 3 years of enrollment.
- Outcome: ctDNA signal detection rate up to 1, 2 or 3 years prior to cancer diagnosis.

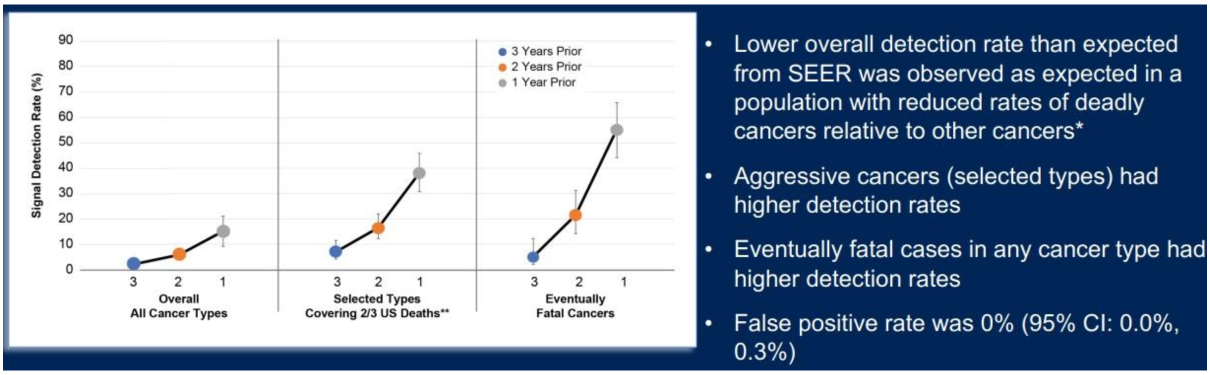

**Question 3a: Do you believe that the ctDNA cancers detected by Galleri have the shortest sojourn time of all breast cancers?**

**Question 3b (if YES to Q3a):**

Galleri test sensitivity for breast cancer is 30.5%, indicating they are ctDNA positive cancers (20.9% of cancers in early stages, 87% of cancers in late stages).

If OMST for all breast cancers is << *INSERT MODE from Q1b*>> years, the predicted sojourn time for the 30.5% of breast cancers with the shortest sojourn time (using a statistical model applied to your answer to question 1c) is << *INSERT VALUE predicted from Q1c*>> years.

**Is this value reflective of your beliefs on the MST of the ctDNA cancers detected by the Galleri test?**

**Question 3c (if NO to Q3a or Q3b):**

Galleri test sensitivity for breast cancer is 30.5%, indicating they are ctDNA positive cancers (20.9% of cancers in early stages, 87% of cancers in late stages).

**If OMST for all breast cancers (ctDNA and non-ctDNA) is << *INSERT MODE from Q1b*>> years, what do you believe is the OMST of ctDNA breast cancers?** (elicited with uncertainty using Chips and Bins)

#### S5. Methods for deriving stage-sojourn times and OMST for ctDNA cancers

To ensure consistency, stage-sojourn times were elicited conditionally on overall mean sojourn time for all cancers and on the proportion of cancers detected in early stage (as opposed to late stage). However, clinically relevant estimates of early-stage mean sojourn time and late-stage mean sojourn time are unconditional and propagate uncertainty appropriately - these have been derived from the elicited quantities as described below.

To derive stage sojourn times for all cancers, for each expert, we derived the ratio between OMST in cancers diagnosed in early stage (OMST_early) and in late stage (OMST_late), where OMST_early was elicited as a point estimate conditioned on OMST in all cancers (OMST_all) in Question 2a, and OMST_late was derived from those two values and the proportion of cancers diagnosed in early stage. We derived the expected (mean) ratio for all experts, assuming that the ratio is independent of OMST_all. We then derived uncertainty in OMST_early and OMST_late, by propagating their ratio through uncertainty in the pooled estimate of OMST_all (elicited in question 1a/1b), ensuring their mean - weighted by the proportion diagnosed in early and late-stages - remained constrained to OMST_all. To derive stage sojourn times for cancers diagnosed in late stage (EMST_late and LMST), we derived the proportion of OMST_late spent in the early stages using the point estimates elicited in question 2b. We derived the expected (mean) proportion for all experts, assuming that the ratio is independent of OMST_late. We then derived uncertainty in EMST_late by propagating this proportion through uncertainty in OMST_late. We derived LMST by subtracting EMST_late from OMST_late

To derive OMST for ctDNA cancers with uncertainty, we first generated predictions for individual experts and pooled them linearly. For each expert, we used the estimates of OMST for ctDNA cancers (OMST_ctDNA) elicited conditioned on the mode OMST_all (question 3) to derive the relative duration of OMST_ctDNA. We then applied the relative duration to uncertain estimates of OMST_all (elicited in question 1a/1b).

#### S6. Elicitation, further results

Table S6.1. Number of experts who provided quantitative estimates for each cancer site.

| Cancer site | Specialists | Generalists | Total |
|---|---|---|---|
| Anus | 1 | 3* | 4 |
| Bladder | 0 | 3* | 3 |
| Breast | 2 | 3 | 5 |
| Cervix | 1 | 3 | 4 |
| Colon/Rectum | 3 | 2 | 5 |
| Gallbladder | 1 | 3* | 4 |
| Head and neck | 2 | 2 | 4 |
| Kidney | 0 | 3* | 3 |
| Liver | 1 | 2 | 3 |
| Lung | 3 | 3 | 6 |
| Lymphoma | 1 | 3 | 4 |
| Melanoma | 0 | 2* | 2 |
| Oesophagus | 2 | 2 | 4 |
| Ovary | 1 | 3 | 4 |
| Pancreas | 3 | 3 | 6 |
| Prostate | 2 | 3 | 5 |
| Sarcoma | 0 | 3* | 3 |
| Stomach | 1 | 3* | 4 |
| Thyroid | 0 | 3* | 3 |
| Urothelial tract | 0 | 2* | 2 |
| Uterus | 0 | 3* | 3 |

* Did not provide quantitative estimates, only indicated similarity to other cancer sites.

**Figure S6.1. Individual estimates of OMST in all cancers**

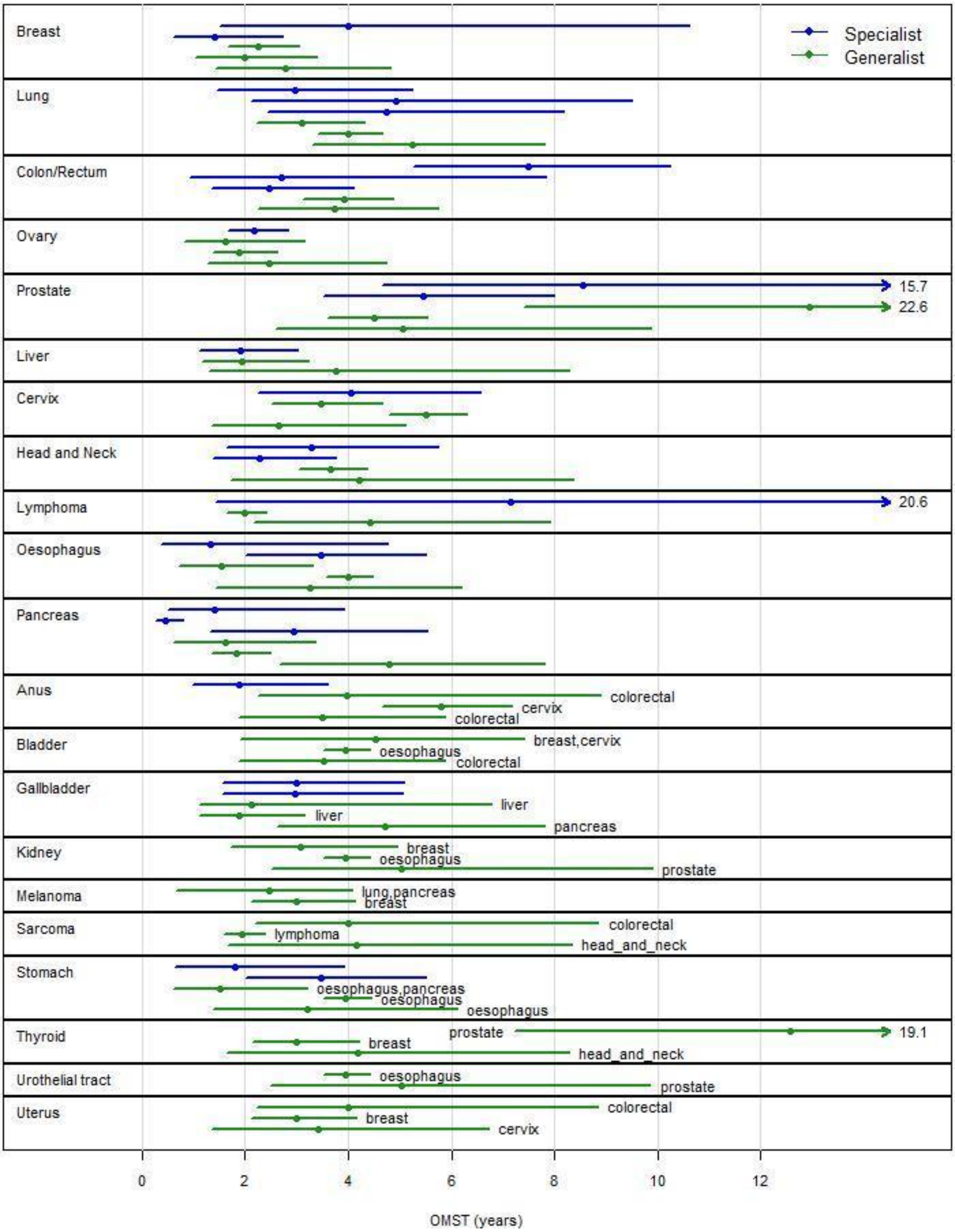

**Figure S6.2. Individual estimates of OMST in all cancers: clinically detected only vs clinically and screen-detected cancers.**

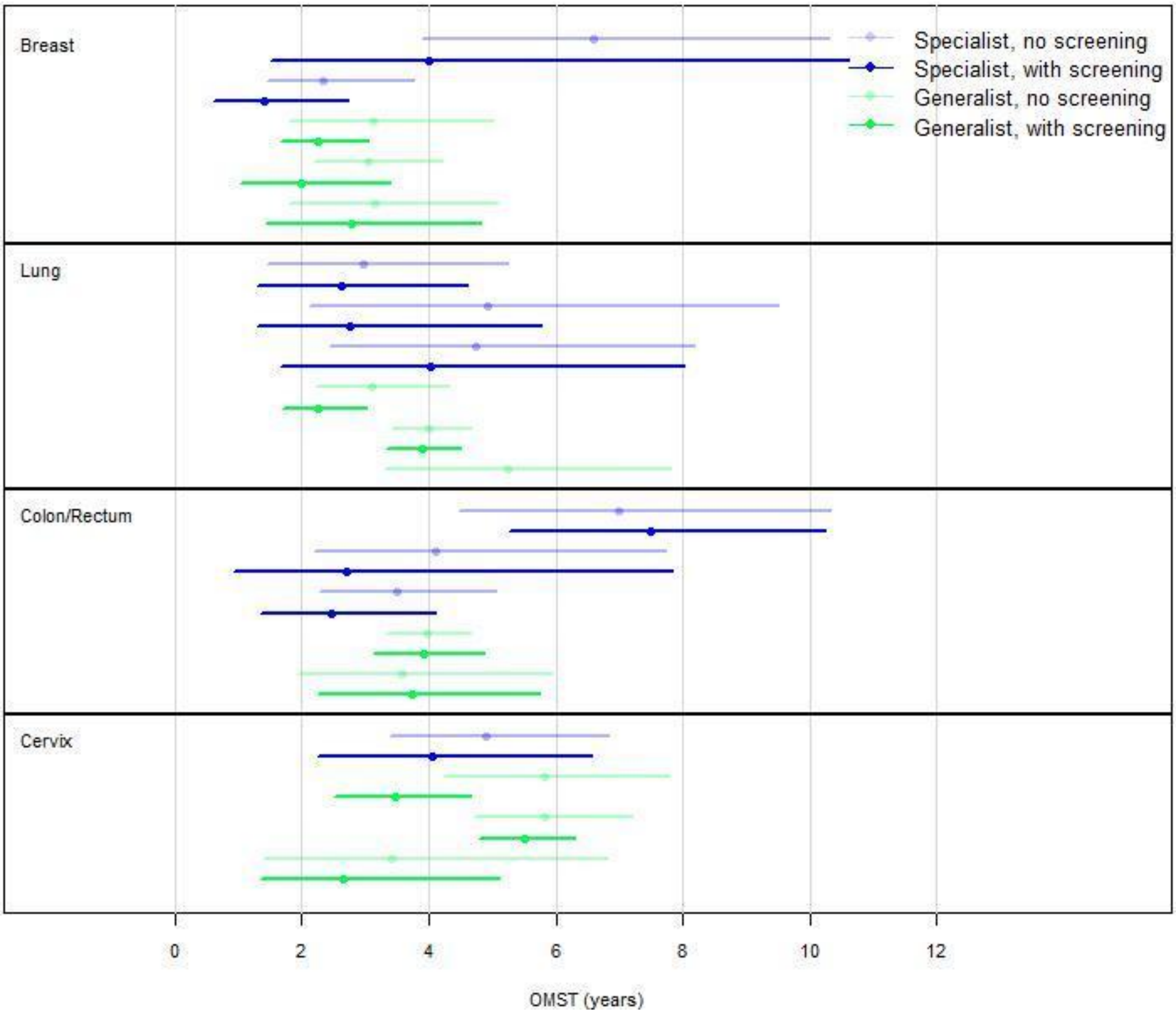

**Table S6.2. Individual estimates of variability of sojourn times within cancer sites.**

| Cancer site | OMST (years) | 25th percentile (years) | Distribution fitted |
|---|---|---|---|
| Breast | 1.4* | 1.0 | lognormal |
| | 6.5* | 3.0 | lognormal |
| | 2.0 | 1.5 | lognormal |
| | 3.25 | 1.75 | lognormal |
| | 2.2 | 1.0 | lognormal |
| Lung | 3.0* | 0.66 | lognormal |
| | 4.5* | 2.0 | lognormal |
| | 4.5* | 3.0 | lognormal |
| | 2.2 | 1.0 | lognormal |
| | 5.25 | 3.0 | lognormal |
| | 3.9 | 3.0 | lognormal |
| Colon/Rectum | 2.5* | 1.5 | lognormal |
| | 2.5* | 1.0 | lognormal |
| | 7.75* | 5.0 | lognormal |
| | 3.75 | 2.5 | lognormal |
| | 4.2 | 3.0 | lognormal |
| Ovary | 2.1* | 0.6 | exponential |
| | 1.8 | 1.0 | lognormal |
| | 2.25 | 1.2 | lognormal |
| | 1.9 | 1.0 | lognormal |
| Prostate | 5.8* | 1.5 | lognormal |
| | 7.5* | 3.0 | lognormal |
| | 4.5 | 4.0 | lognormal |
| | 4.5 | 2.5 | lognormal |
| | 12 | 6.0 | lognormal |
| Liver | 1.8* | 0.7 | lognormal |
| | 2.0 | 1.0 | lognormal |
| | 3.0 | 1.5 | lognormal |
| Cervix | 4.75* | 4.0 | lognormal |
| | 3.8 | 3.0 | lognormal |
| | 5.5 | 4.0 | lognormal |
| | 3.25 | 1.5 | lognormal |
| Head and neck | 3.2* | 1.5 | lognormal |
| | 2.2* | 1.0 | lognormal |
| | 3.5 | 3.0 | lognormal |
| | 4.0 | 2.0 | lognormal |
| Lymphoma | 3.0* | 2.0 | lognormal |
| | 2.0 | 10 | lognormal |
| | 4.25 | 2.4 | lognormal |
| Oesophagus | 1.0* | 0.5 | lognormal |
| | 3.5* | 2.0. | lognormal |
| | 1.2 | 0.7 | lognormal |
| | 4.0 | 3.0 | lognormal |
| | 3.0 | 1.75 | lognormal |
| Pancreas | 0.4* | 0.2 | lognormal |
| | 3.0* | 1.5 | lognormal |
| | 1.2* | 0.4 | lognormal |
| | 1.6 | 1.0 | lognormal |
| | 1.9 | 1.0 | lognormal |
| | 4.75 | 3.0 | lognormal |
| Anus | 1.6* | 10 | lognormal |
| Gallbladder | 3.0* | 1.5 | lognormal |

| | | | |
|---|---|---|---|
| | 2.8* | 1.0. | lognormal |
| Oesophagus | 3.5* | 2.0 | lognormal |
| | 1.8* | 0.8 | lognormal |

Green/red indicate the distribution was/wasn't validated in question 3. While indicates experts were not asked to validate as they did not believe that ctDNA cancers were cancers with the shortest OMST within the cancer site.
* Estimate provided by specialist

**Figure S6.3. Individual estimates of conditional* stage-sojourn times in all cancers.**

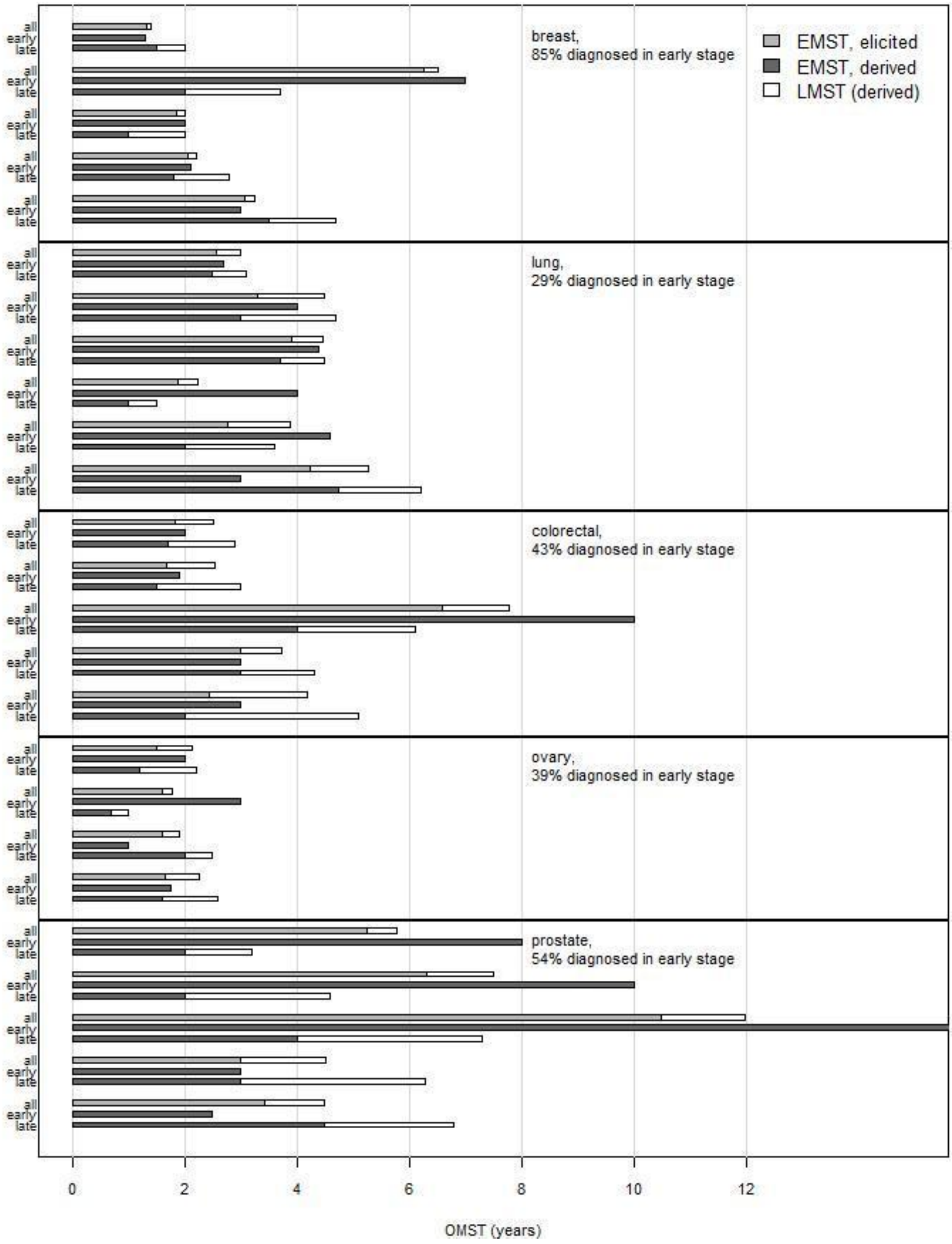

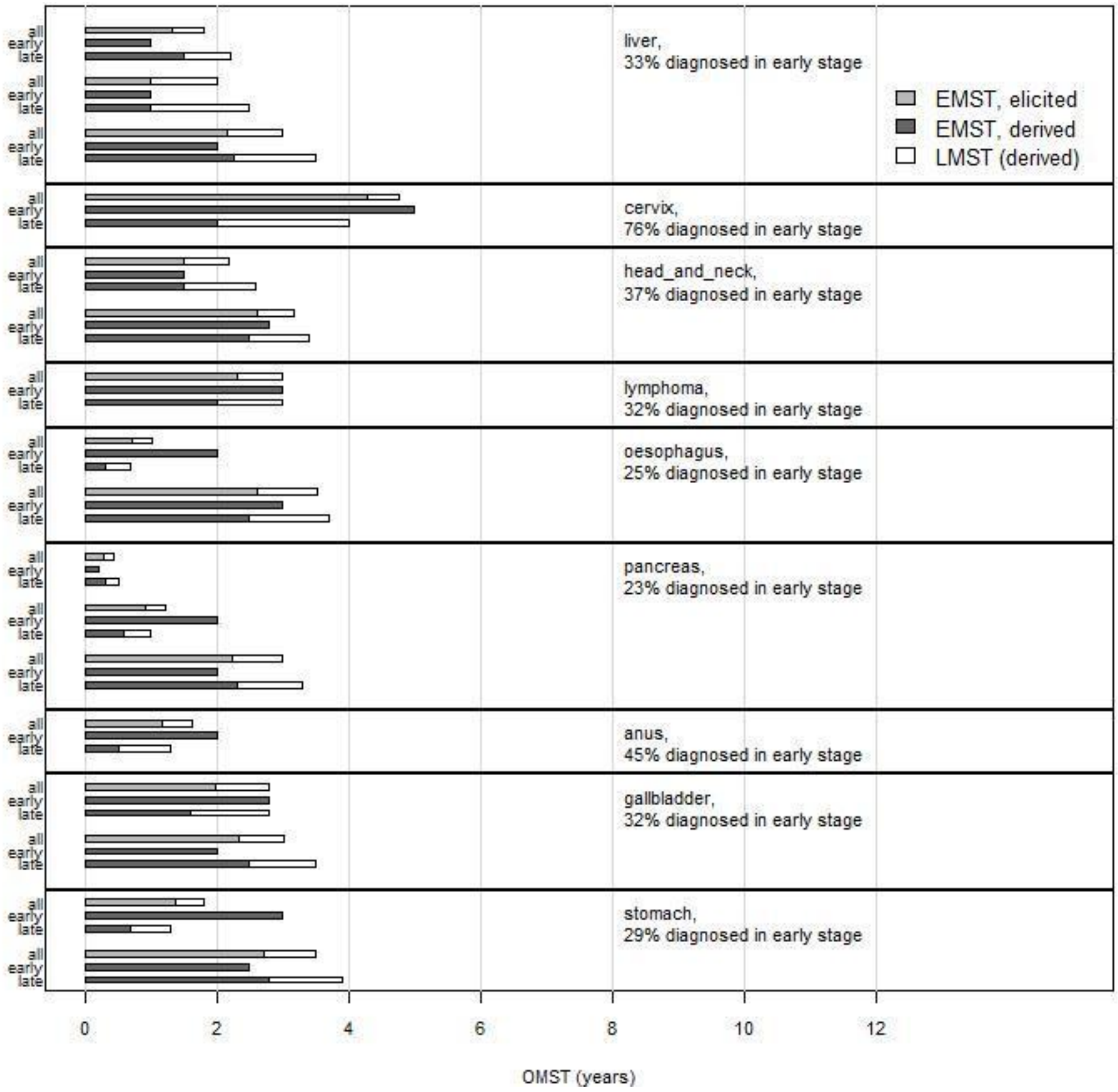


* These estimates were elicited conditioned on the individual expert's OMST point estimate. The figure shows that in 68% (28/41) of the estimates, EMST is shorter in cancers diagnosed in late stages than cancers diagnosed in early stages, indicating that cancers diagnosed late tend to progress faster. Furthermore, in 85% (35/41) of the estimates for cancers diagnosed in late stage, LMST is shorter than EMST.

**Table S6.3. Individual estimates of OMST in ctDNA cancers.**

| Cancer site | Galleri test sensitivity across all stages | OMST (years) in all cancers | OMST (years) in ctDNA cancers: point estimate/median (95% CrI) |
|---|---|---|---|
| Breast | 30.5% | 1.4* | 0.7 (0.6, 0.9) |
| | | 6.5* | 5.6 (4.5, 6.9) |
| | | 2 | 0.3 |
| | | 2.2 | 1.6 (1.2, 2) |
| | | 3.25 | 0.6 |
| Lung | 74.8% | 3* | 1.6 |
| | | 4.5* | 2.4 |
| | | 4.5* | 2.8 (1.3, 5.3) |
| | | 2.2 | 1.2 |
| | | 3.9 | 4 (3.6, 4.4) |
| | | 5.25 | 2.8 |
| Colon/Rectum | 82.0% | 2.5* | 1.7 (1.1, 2.6) |
| | | 2.5* | 2.5 (2, 3.1) |
| | | 7.75* | 7.4 (6, 9.3) |
| | | 3.75 | 2.3 |
| | | 4.2 | 2.6 |
| Ovary | 83.1% | 2.1* | 1.5 (1.1, 1.9) |
| | | 1.8 | 1.2 |
| | | 1.9 | 1.2 |
| | | 2.25 | 1.4 |
| Prostate | 11.2% | 5.8* | 1.2 (0.9, 1.4) |
| | | 7.5* | 8 (6.7, 9.6) |
| | | 12 | 7.8 (4.3, 12.9) |
| | | 4.5 | 0.3 |
| | | 4.5 | 0.3 |
| Liver | 93.5% | 1.8* | 1.8 (1.5, 2.1) |
| | | 2 | 1.6 |
| | | 3 | 2.4 |
| Cervix | 80.0% | 4.75* | 3.9 (2, 6.6) |
| | | 3.25 | 1.9 |

| | | | |
|---|---|---|---|
| | | 3.8 | 2.3 |
| | | 5.5 | 3.3 |
| Head and Neck | 85.7% | 2.2* | 2 (1.9, 2.2) |
| | | 3.2* | 2.2 |
| | | 3.5 | 2.4 |
| | | 4 | 2.7 |
| Lymphoma | 56.3% | 3* | 1.9 (0.7, 3.8) |
| | | 2 | 0.7 |
| | | 4.25 | 1.5 |
| Oesophagus | 85.0% | 1* | 0.7 |
| | | 3.5* | 2.3 |
| | | 1.2 | 0.8 |
| | | 3 | 2 |
| | | 4 | 2.7 |
| Pancreas | 83.7% | 1.2* | 0.8 (0.4, 1.5) |
| | | 3* | 1.9 |
| | | 1.6 | 1 |
| | | 1.9 | 1.2 |
| | | 4.75 | 3.1 |
| Anus | 81.8% | 1.6* | 1.4 (1.1, 2) |
| Gallbladder | 70.6% | 2.8* | 2.6 (2.2, 3) |
| | | 3* | 1.5 |
| Stomach | 66.7% | 1.8* | 1.1 (0.6, 1.7) |
| | | 3.5* | 1.6 |

**Figure S6.4. Individual estimates of OMST in ctDNA cancers, by cancer site (Galleri test sensitivity).**

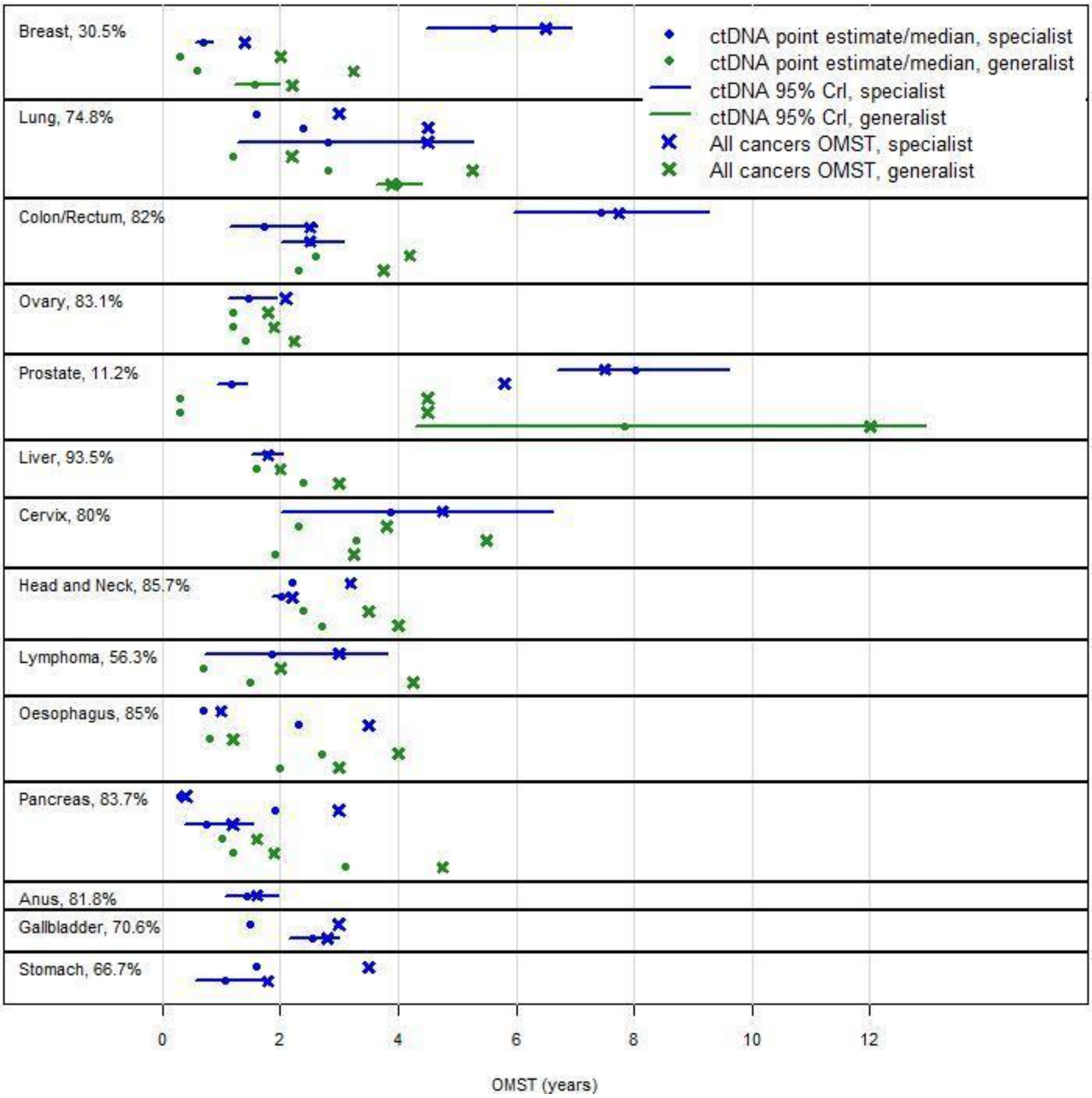

**Figure S6.5. Derived OMST for ctDNA, by expert and cancer site.**

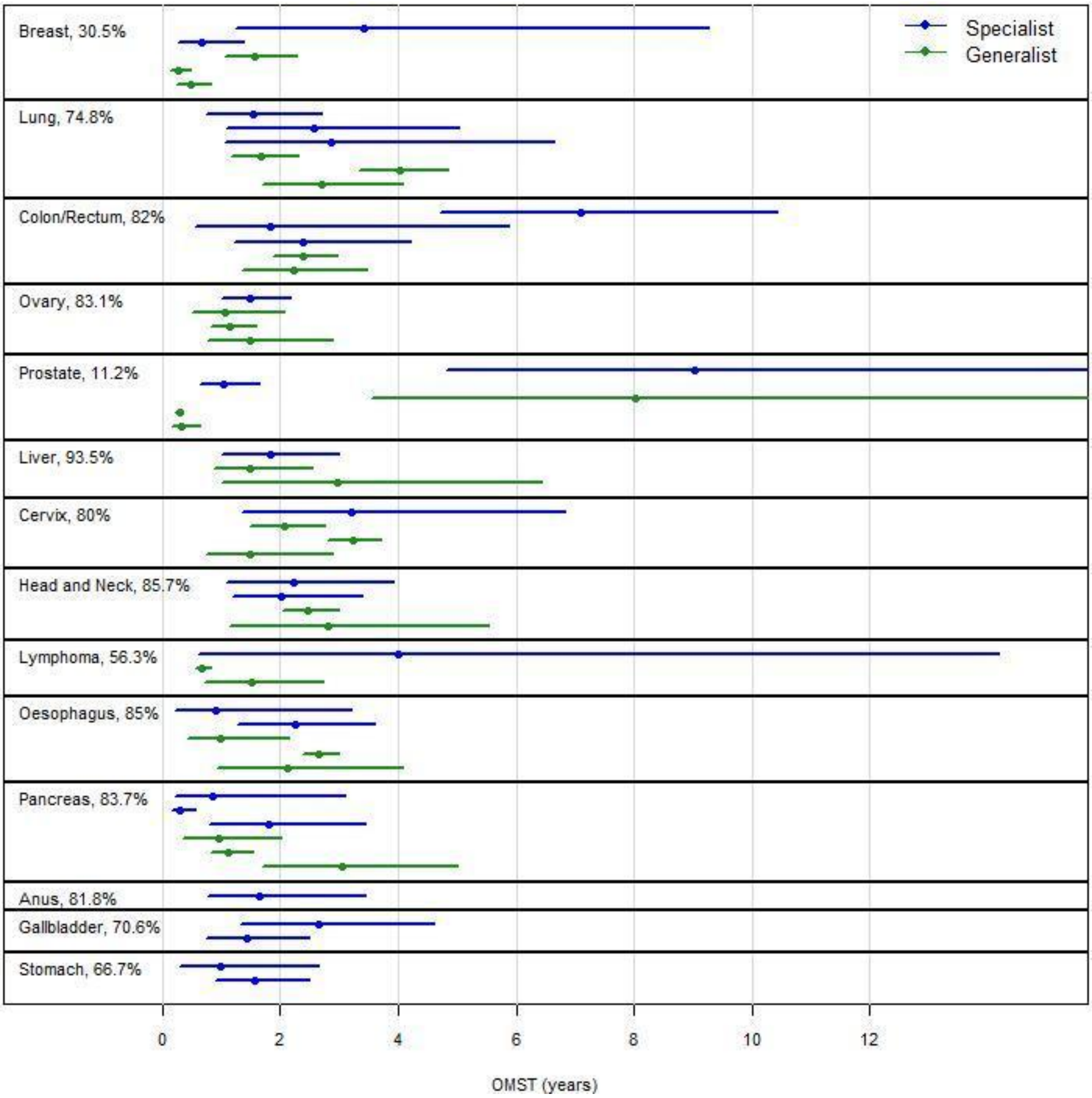

**Table S6.4. All estimated and derived sojourn times**

| Cancer site | All cancers | | | | | ctDNA cancers |
|---|---|---|---|---|---|---|
| | OMST | OMST/early stage-MST, detected early | OMST, detected late | early stage-MST, detected late | late stage-MST | OMST |
| Breast | 2.3 (0.9, 7.1) | 2.3 (0.9, 7) | 2.3 (0.9, 7.1) | 1.5 (0.6, 4.5) | 0.8 (0.3, 2.6) | 0.8 (0.2, 6.2) |
| Lung | 4 (2, 7.8) | 4.5 (2.2, 8.8) | 3.8 (1.9, 7.4) | 2.7 (1.3, 5.3) | 1.1 (0.5, 2.2) | 2.5 (1, 5) |
| Colon/Rectum | 3.8’l(‘1.4, 9.1) | 3.4 (1.2, 8.3) | 4 (1.4, 9.7) | 2.3 (0.8, 5.5) | 1.7 (0.6, 4.2) | 2.5 (1, 9) |
| Ovary | 2 (1.1, 3.8) | 2.3 (1.2, 4.4) | 1.9 (1, 3.5) | 1.2 (0.6, 2.3) | 0.6 (0.3, 1.2) | 1.3 (0.7, 2.4) |
| Prostate | 6 (3.2, 18) | 7.2 (3.8, 21.4) | 4.6 (2.5, 13.9) | 2.6 (1.4, 7.6) | 2.1 (1.1, 6.3) | 1.1 (0.2, 15.2) |
| Liver | 2.2 (1.2, 6.9) | 1.2 (0.7, 4) | 2.6 (1.4, 8.3) | 1.5 (0.8, 4.8) | 1.1 (0.6, 3.5) | 1.9 (1, 5.4) |
| Cervix | 3.8 (1.7, 6.2) | 4 (1.8, 6.5) | 3.2 (1.5, 5.2) | 1.6 (0.7, 2.6) | 1.6 (0.7, 2.6) | 2.4 (1, 5.3) |
| Head and Neck | 3.4 (1.5, 6.8) | 2.7 (1.2, 5.4) | 3.8 (1.7, 7.7) | 2.5 (1.1, 5) | 1.3 (0.6, 2.6) | 2.4 (1.2, 4.7) |
| Lymphoma | 3.8 (1.7, 16.2) | 3.8 (1.7, 16.2) | 3.8 (1.7, 16.2) | 2.5 (1.1, 10.8) | 1.3 (0.6, 5.4) | 1.4 (0.6, 12) |
| Oesophagus | 3 (0.6, 5.3) | 4.5 (0.9, 8) | 2.5 (0.5, 4.4) | 1.4 (0.3, 2.4) | 1.1 (0.2, 2) | 1.9 (0.4, 3.6) |
| Pancreas | 1.9 (0.3, 6.4) | 1.9 (0.3, 6.4) | 1.9 (0.3, 6.4) | 1.2 (0.2, 4.1) | 0.7 (0.1, 2.4) | 1.2 (0.2, 4.2) |
| Anus | 3.9 (1.2, 7.4) | 4.8 (1.5, 9.2) | 3.1 (1, 6) | 1.2 (0.4, 2.3) | 1.9 (0.6, 3.7) | 1.7 (0.8, 3.5) |
| Bladder | 4 (2.1, 6.8) | NA | NA | NA | NA | NA |
| Gallbladder | 2.8 (1.3, 6.7) | 2.4 (1.1, 5.7) | 3 (1.4, 7.2) | 1.9 (0.9, 4.7) | 1.1 (0.5, 2.6) | 2 (0.9, 4.4) |
| Kidney | 4 (2.1, 8.4) | NA | NA | NA | NA | NA |
| Melanoma | 2.9 (0.9, 4.2) | NA | NA | NA | NA | NA |
| Sarcoma | 3.4 (1.7, 8.3) | NA | NA | NA | NA | NA |
| Stomach | 3 (0.8, 5.3) | 3.9 (1.1, 6.9) | 2.6 (0.7, 4.6) | 1.7 (0.5, 2.9) | 1 (0.3, 1.7) | 1.4 (0.4, 2.6) |
| Thyroid | 4.3 (2.1, 19.5) | NA | NA | NA | NA | NA |
| Urothelial tract | 4.1 (2.9, 8.9) | NA | NA | NA | NA | NA |
| Uterus | 3.4 (1.8, 7.7) | NA | NA | NA | NA | NA |
